# Early, Late, & Self-Selected Time-Restricted Eating: Impact on Hepatic Fat, Liver Health, & Fecal Microbiota in Adults with Overweight or Obesity

**DOI:** 10.64898/2026.01.17.26344338

**Authors:** Manuel Dote-Montero, Antonio Clavero-Jimeno, Adrián Cortés-Martín, Amaya Lopez-Pascual, Elisa Merchan-Ramirez, Alba Camacho-Cardenosa, Mara Concepción, Maddi Osés, Alejandro López-Vázquez, Francisco J. Amaro-Gahete, Juan J. Martin-Olmedo, Lucas Jurado-Fasoli, Alejandro De-la-O, Patricia V. García Pérez, Julio Gálvez, Alba Rodriguez-Nogales, Federico Garcia, Carmen Mbongo Habimana, Marcos Jiménez Vázquez, Víctor Manuel Alfaro-Magallanes, Matías A Avila, Jose L. Martín-Rodríguez, Rafael Cabeza, Manuel Muñoz-Torres, Idoia Labayen, Jonatan R. Ruiz

## Abstract

**Background and aims:** The optimal eating window for time-restricted eating (TRE) remains unclear. We investigated the effects of 8-hour TRE combined with usual care (UC, a Mediterranean diet-based education program), versus UC alone over 12 weeks on hepatic fat fraction, liver health markers, and fecal microbiota in adults with overweight or obesity.

**Methods:** In this multicenter randomized trial, participants (50% women) were assigned to UC (n=49), early TRE (n=49), late TRE (n=52), or self-selected TRE (n=47). Hepatic fat fraction was assessed by MRI; liver markers included elastography-based parameters, liver enzymes, and circulating biomarkers. Fecal microbiota was analyzed by 16S rRNA gene sequencing.

**Results:** Hepatic fat fraction decreased significantly within the three TRE groups (all *P*≤0.02), but no between-group differences were observed when comparing early TRE (mean difference [MD]: -0.4%; *P*=0.95), late TRE (MD: -1.5%; *P*=0.15), and self-selected TRE groups (MD: -0.7%; *P*=0.77) with the UC group, or among the TRE groups themselves (all *P*≥0.41). Similarly, no between-group differences were found in liver health markers and fecal microbiota. Participants with metabolic dysfunction-associated steatotic liver disease at baseline as well as those achieving ≥5% weight loss had greater reductions in hepatic fat fraction than those who did not (MD: -2.7 and -2.6%; respectively, both *P*<0.001). A higher proportion of participants in the TRE groups achieved ≥5% weight loss compared with UC (41–44% vs 16%; P=0.001).

**Conclusion:** These findings suggest that the timing of the eating window in TRE may not impact liver fat or microbiota composition beyond the effects of weight loss, though the study was not powered for secondary outcomes.

The study was registered on ClinicalTrials.gov (identifier: NCT05310721)

## INTRODUCTION

Obesity is a major public health challenge,^1^ increasing the risk of metabolic dysfunction-associated steatotic liver disease (MASLD).^2^ MASLD is characterized by a prolonged accumulation of lipids in the liver and is tightly linked to type 2 diabetes and cardiovascular disease,^3,4^ placing a substantial burden on healthcare systems.^5^ Although calorie restriction effectively improves hepatic steatosis and metabolic health,^6–8^ its long-term adherence remains challenging.^9^

Time-restricted eating (TRE) has emerged as a promising dietary approach that limits food intake to a predefined eating window of ≤10 hours, followed by a fasting period ≥14 hours.^10–12^ Notably, our group^13^ and others have shown that TRE is well tolerated, achieves high adherence rates, and leads to minimal adverse effects, resulting in modest body weight loss and slight improvements of cardiometabolic health in individuals with overweight or obesity.^11,12,14^ Whether the timing of the eating window influences TRE’s effectiveness on hepatic fat fraction and liver health markers remains uncertain.^10^

Few studies have investigated the TRE’s effects on hepatic fat fraction using magnetic resonance imaging (MRI) or MRI combined with magnetic resonance spectroscopy,^15–17^ both consolidated, non-invasive gold-standard methods for measuring hepatic fat fraction.^18^ Overall, these studies suggest that TRE reduces hepatic fat fraction while improving liver function, but limitations such as absence of control group, short duration of the intervention, or exclusive focus on early TRE schedules, prevent definitive conclusions. We recently reported that, after a 12-week intervention, early TRE (eating window from ∼09:45 to ∼17:30), late TRE (∼14:20 to ∼21:30), and self-selected TRE (∼12:20 to ∼20:00) combined with usual care (UC), a Mediterranean diet-based education program, significantly reduced visceral adipose tissue as measured by MRI; however these reductions did not differ from UC alone (∼08:30 to ∼22:00).^13^ However, early TRE significantly lowered fasting and nocturnal glucose levels compared to UC and the other TRE schedules, and also decreased subcutaneous abdominal adipose tissue relative to UC.^13^ The liver exhibits robust circadian rhythm, which regulates the expression and activity of enzymes involved in glucose and lipid metabolism peaking in the morning in humans.^19^ Therefore, it is biologically plausible that early TRE may elicit a greater impact on hepatic fat fraction and liver health markers.

Additionally, gut-liver axis dysfunction has been identified as a contributing factor to MASLD, with negative alterations in gut microbiota composition and function leading to increased lipid absorption and triggering a proinflammatory cascade that exacerbates hepatic inflammation through dysregulated hepatokines and cytokines secretion.^20^ Thus, understanding the potential interactions between TRE, gut microbiota composition, and hepatic fat fraction may provide insights into novel therapeutic approaches for MASLD.

In this randomized controlled trial (RCT), we investigated the effects of three distinct TRE schedules, an 8-hour eating window in the early part of the day (early TRE), an 8-hour window later in the day (late TRE), and a participant-selected eating window (self-selected TRE), combined with UC, versus UC alone over 12 weeks. This study focused on changes in hepatic fat fraction measured by MRI, quantitative elastography-based liver markers (including stiffness, attenuation, and viscosity), liver enzymes, circulating liver-related adipo- and hepatokines, and fecal microbiota composition in men and women with overweight or obesity.

## METHODS

### Study design

This study is a secondary analysis of a multicenter, parallel-group, investigator-initiated RCT. Comprehensive details on the study rationale, design, and methodology are available in the published protocol.^21^ The trial received approval from the relevant regulatory bodies and ethics committees in Spain, including the *Servicio Andaluz de Salud*, the *Comité Ético de Investigación Provincial de Granada* (ref. 1/22), and the *Comité Ético de Investigación Clínica de Navarra* (PI_2021/119). The study was registered on ClinicalTrials.gov (identifier: NCT05310721) and conducted in accordance with the Consolidated Standards of Reporting Trials (CONSORT) guidelines for randomized clinical trials.^22^ Written informed consent was obtained from all participants prior to enrollment.

### Participants and Eligibility Criteria

Men and women aged 30 to 60 years with overweight or obesity (body mass index [BMI] between 25.0 and 40.0 kg/m²) were eligible for inclusion if they also met the following criteria: abdominal obesity (waist circumference ≥95 cm for men and ≥82 cm for women), stable body weight (±3%) for ≥3 months, <150 min/week of moderate-to-vigorous physical activity, a habitual eating window ≥12 hours, and at least one cardiometabolic risk factor associated with metabolic syndrome.^21^ Key exclusion criteria included ongoing participation in weight-loss programs, the presence of cardiovascular or chronic diseases (e.g., recent myocardial infarction, hemorrhagic stroke, Cushing’s syndrome, adrenocortical insufficiency, or type 1 or 2 diabetes), pregnancy or breastfeeding, and current night-shift work. Further details regarding the eligibility criteria and screening procedures are described elsewhere.^21^ Participants did not receive financial compensation.

### Study Recruitment, Enrollment, and Randomization

Participants were enrolled in nine consecutives waves of 10–13 individuals per center between April 11, 2022, and December 5, 2022, with final study procedures completed by March 6, 2023. Recruitment was carried out through newspaper advertisements and referrals from the Endocrinology and Nutrition departments at *Hospital Universitario Clínico San Cecilio and Hospital Virgen de las Nieves* in Granada, as well as the *Hospital Universitario de Navarra* in Pamplona. Prior to enrollment, prospective participants underwent clinical and physical examinations to confirm eligibility. Those who met the inclusion criteria proceeded to baseline assessments.

Following baseline assessments, participants were randomized to one of four intervention arms (UC, early TRE, late TRE, or self-selected TRE) using a stratified, permuted block design. Randomization was stratified by study site and sex, with variable block sizes (4 and 8). Separate randomization lists were generated for each site and stratum. Within each block, participants were randomly assigned in equal proportions to one of the four study groups using a 1:1:1:1 allocation ratio under a parallel-group design.

Personnel responsible for acquiring and analyzing MRI-derived hepatic fat fraction, ultrasound-based outcomes, and for analyzing fasting blood samples and fecal microbiota composition were blinded to group allocation. In contrast, staff involved in delivering the intervention and performing other assessments were not blinded (open-label design).

### Study Assessments and Outcomes

#### Hepatic fat fraction

Assessments were conducted at baseline and after the 12-week intervention. Hepatic fat fraction was measured using MRI (Siemens 3T Magnetom Vida, Siemens Healthineers, Erlangen, Germany) under fasting conditions (minimum 4–5 hours). Stages of hepatic steatosis were defined as follows: S0: no steatosis (hepatic fat fraction <5%); S1: mild steatosis (hepatic fat fraction ≥5% and <15%]; S2: moderate steatosis (hepatic fat fraction ≥15% and <25%); S3: severe steatosis (hepatic fat fraction ≥ 25%).^23^

#### Elastography

Liver stiffness and viscosity were assessed using shear wave elastography and shear wave dispersion, respectively, while liver steatosis was assessed using the attenuation imaging coefficient (ATI). All ultrasound-based assessments were conducted using the Aplio i800 system (Canon Medical Systems Corporation, Tochigi, Japan) under fasting conditions (minimum 4–5 hours).

#### Liver enzymes

Liver enzyme levels, including alanine aminotransferase (ALT), γ–glutamyltransferase (GGT), and alkaline phosphatase (ALP), were measured from serum samples collected after a 10–12-hour overnight fast using an AU5800 automated analyzer (Beckman Coulter Inc., CA, USA).

#### Anthropometric

Body weight and height were measured using a calibrated electronic column scale and stadiometer (Seca model 799, Hamburg, Germany).^24^ BMI was calculated as weight in kilograms divided by height in meters squared. Waist and hip circumferences were measured following the International Society for the Advancement of Kinanthropometry (ISAK) protocols.^24^

#### Adipokines, hepatokines, and adipo-hepatokines

Adipokines, hepatokines, and adipo-hepatokines were quantified using commercial Enzyme-Linked Immunosorbent Assay (ELISA) kits (R&D Systems, MN, USA), following the manufacturer’s specifications. The sample type (serum or plasma) and the specific ELISA kits were used for measurements as detailed: adiponectin (DY1065), interleukin-6 (IL-6; HS600C), angiopoietin-like 3 (ANGPTL3; DY3829), clusterin (DY5874), sex hormone-binding globulin (SHBG; DY2656), dipeptidyl peptidase IV (DPPIV; DY1180), and retinol-binding protein 4 (RBP4; DY3378) were measured from serum, whereas omentin (DY4254), resistin (DY1359), β-Klotho (DY5889), follistatin (DY669), angiopoietin-like 4 (ANGPTL4; DY3485), and growth differentiation factor 15 (GDF15; DY957) were measured from plasma.

#### Dietary intake

Dietary intake was assessed using three non-consecutive 24-hour dietary recalls (two on working days and one on a nonworking day) administered through face-to-face or online interviews conducted by qualified and trained research nutritionists. Energy intake, macronutrient intake, and alcohol intake were calculated from the 24-hour dietary recalls using the EasyDiet-Programa de Gestión de la Consulta® de la Academia Española de Nutrición y Dietética (Biocentury, S.L.U. 2016), computer software supported by the Spanish Association of Dietitians and Nutritionists.

#### Insulin resistance

Venous blood samples were collected and stored at −80°C for the analysis of fasting glucose and insulin using automated analyzers as previously described.^21^ Then, the homeostasis model assessment of insulin resistance (HOMA-IR) was calculated.^25^ Prior to sample collection, participants were instructed to fast for approximately 10 hours, abstain from alcohol and diuretics for 24 hours, avoid stimulants such as caffeine for 12 hours, and refrain from moderate physical activity for 24 hours or vigorous physical activity for 48 hours.

#### Fecal microbiota

A total of 182 participants provided fecal samples in sterile containers before and during the last two weeks of the intervention for fecal microbiota analysis through 16S rRNA amplicon sequencing. Notably, due to sequencing technical issues and low-quality data, the final analysis included gut microbiota data from 138 participants at both time points of the intervention. These participants were evenly distributed across the different intervention groups (UC: n = 35; early TRE: n = 36; late TRE: n = 36; self-selected TRE: n = 31). Samples were self-collected either the evening before or on the same morning they were delivered to the laboratory. The samples collected the evening before were stored in the home freezer prior to being transported in cooler containers to the laboratory. Upon arrival, all fecal samples were stored at -80 °C until further analysis. Bacterial DNA was extracted from the samples using the QIAamp PowerFecal Pro DNA Kit (Qiagen, Hilden, Germany) following the manufacturer’s instructions. Library preparation was conducted following the 16S Metagenomic Sequencing Library Preparation Illumina protocol. 16S bacterial rRNA gene was amplified using primers targeting the V3-V4 hypervariable region (341F and 805R primers). Sequencing was performed on a NextSeq1000-Illumina platform (Illumina Inc., San Diego, CA, USA) with a 2 x 300 bp paired-end read length at the Microbiology Sequencing Service of the *Hospital Universitario Clínico San Cecilio* (Granada, Spain). Bioinformatic analysis of the sequencing data was carried out using the Quantitative Insights Into Microbial Ecology (QIIME2) (v.2024.10) workflow.^26^ Briefly, demultiplexed sequences were processed to remove primers sequences using the q2-cutadapt plugin. Denoising to eliminate low-quality bases, dereplication, merging of denoised paired-end reads, chimera removal, and generation of the Amplicon Sequence Variants (ASVs) table were carried out using the DADA2 algorithm.^27^ After denoising, an average of 303,426.47 (± 183,940.94, standard deviation) sequences per sample were retained. ASVs were filtered to remove those that appeared in less than 10% of the samples. Taxonomic classification was assigned using a pre-trained classifier based on the SILVA 138.1 SSU reference database,^28^ considering the genus-level taxonomy as the highest classification category. Alpha-diversity indices (Shannon entropy, Simpson index, observed features, and Pielou evenness) and beta-diversity (Bray-Curtis dissimilarity) were calculated using the q2-diversity plugin, with a sampling depth of 38,500 reads per sample. Microbial functions were predicted by Phylogenetic Investigation of Communities by Reconstruction of Unobserved States (PICRUSt2) (v.2.4.2).^29^ Absolute counts for taxa and microbial functions were transformed into relative abundance to normalize the data.

### Study Intervention and Control Condition

Participants assigned to the UC group maintained their habitual eating schedule (≥12-hour eating window) and received a healthy lifestyle educational program,^21^ based on the Mediterranean dietary pattern ^30^ and aligned with the World Health Organization’s physical activity recommendations.^31^ All participants, including those in the TRE groups, attended twice-monthly sessions led by experienced dietitians. These meetings addressed six key topics: (i) healthy lifestyle practices based on Mediterranean diet and physical activity guidelines; (ii) meal planning and organization; (iii) hunger and satiety regulation; (iv) interpreting food labels; (v) nutritional myths; and (vi) healthy snack selection. Each session concluded with a group discussion of individual challenges, allowing for personalized guidance and peer support.

Participants in the early TRE group selected an 8-hour eating window that began no later than 10:00, while those in the late TRE group chose an 8-hour window starting no earlier than 13:00. The self-selected TRE group established their preferred 8-hour eating window prior to the start of the intervention. All TRE participants were instructed to maintain the same eating window throughout the 12-week intervention and to adhere to it daily (i.e., seven days per week). Consumption of calorie-containing foods or beverages outside the eating window was not permitted; only water, black coffee, and unsweetened tea (free of sugar or artificial sweeteners) were allowed. All participants, including those in the UC group, recorded the timing of their first and last meals each day throughout the intervention using a customized mobile application (EXTREME: com.nnbi.app_extreme, NNBi2020 S.L., Navarra, Spain).

Given that Spain is known for having one of the latest dinner times in Europe—typically around 22:00, compared to 21:00 in Italy, 20:00 in France, 19:00 in Germany, and 18:00 in Sweden^32^—we allowed early TRE participants to choose an 8-hour window that started no later than 10:00, enabling a dinner as late as 18:00. While this may be considered a typical dinner time in other countries, it remains early by Spanish standards.

### Statistical analysis

Sample size and power calculations for the trial were based on the primary outcome (changes in visceral adipose tissue) of the original study, as described elsewhere.^13^ Since the present work focuses exclusively on secondary outcomes, no separate *a priori* sample size or power estimations were conducted for these analyses. The Shapiro-Wilk test was used to assess the normality of data distributions. Owing to skewed distribution, adipokines, hepatokines, and adipo-hepatokines were log10-transformed to normalize their distributions for statistical analyses; however, results are presented in the original units to facilitate interpretation. Intervention impact on secondary outcomes (i.e., liver parameters, adipokines, hepatokines, adipo-hepatokines, dietary intake, and alpha diversity metrics from fecal microbiota) in response to the 12-week intervention were evaluated using repeated-measures linear mixed-effects models, incorporating random effects for study site.^33^ Changes in individual outcomes were modeled as a function of intervention group, site, time, and their interaction terms. Sex was included as a covariate in all models. Analyses were performed under the intention-to-treat principle using restricted maximum likelihood estimation, assuming missing data were missing at random. To adjust for multiple comparisons, Tukey’s method was applied and reported *P* values are Tukey-adjusted. To examine differences in MASLD remission (defined as hepatic fat fraction <5%) across intervention groups among participants with MASLD at baseline, Fisher’s exact test was used.

Regarding fecal microbiota analyses, to examine differences in baseline fecal microbiota between groups, a multivariate analysis of variance (MANOVA) was performed after principal component analysis (PCA) of baseline relative abundances of taxa. The Kruskal-Wallis test, followed by Dunn’s *post-hoc* test, was used to compare baseline alpha diversity indices. Differences in beta diversity, based on the Bray-Curtis dissimilarity, were tested using permutational multivariate analysis of variance (PERMANOVA) with 999 permutations. *P* values for both alpha and beta diversity were adjusted using the Bonferroni correction for multiple comparisons. Beta diversity was visualized using principal coordinates analysis (PCoA) plots in all analyses. Within-group differences (i.e., differences between preintervention [PRE] and postintervention [POST] timepoints for each group) were evaluated using the paired t-test or the Wilcoxon signed-rank test for alpha diversity metrics, depending on whether the data were normally distributed or not, and PERMANOVA was used for beta diversity. Linear discriminant analysis effect size (LEfSe)^34^ was performed to identify characteristic biomarkers of bacterial taxa and metabolic pathways associated with the intervention within each group by comparing time points (e.g., UC PRE versus UC POST, early TRE PRE versus early TRE POST, etc.). The linear discriminant analysis (LDA) score was set at 2.0. Changes produced in beta diversity during the intervention were tested using PERMANOVA, evaluating the interaction between group and assessment time. Microbiome Multivariable Associations with Lineal Models (MaAsLin3)^35^ were conducted to evaluate changes in taxonomic composition, both in terms of abundance and prevalence, considering the interaction between group and assessment time, adjusting by sex, and including participant ID as a random effect. The UC group was used as the reference group. Multiple testing correction was applied using the Benjamini-Hochberg False Discovery Rate (FDR).^36^ Furthermore, we conducted sensitivity and *post-hoc* analyses by repeating the linear mixed-effects models for all outcomes, stratified by sex (i.e., separately for men and women).

Then, we examined the association between changes in hepatic fat fraction and changes in adipokines, hepatokines, adipo-hepatokines, insulin resistance, and fecal microbiota diversity over time using repeated measures correlation analysis, a method that assesses within-individual associations across multiple time points.^37^ Pearson’s chi-square test was conducted to examine the association between the intervention group and the achievement of a clinically meaningful weight loss (≥5%). We then conducted exploratory analyses comparing changes in hepatic fat fraction and liver-related biomarkers between participants who achieved clinically meaningful weight loss (≥5%; so-called responders) and those who did not (<5%; so-called non-responders). Between-group differences over time were evaluated using repeated-measures linear mixed-effects models that included a group × time interaction term, a random intercept for participant ID, and sex as a covariate. *P* values were corrected for multiple comparisons across outcomes using the Benjamini-Hochberg FDR method.

All statistical analyses were conducted using R (v.4.4.2; R Foundation for Statistical Computing, Vienna, Austria; https://cran.r-project.org/), with linear mixed-effects models fitted using the *lme4 package (v.1.1-36)*, and graphical representations were generated using the *ggplot2 package (v.3.5.1)*. PERMANOVA was performed using the *vegan package (v.2.6.10)* and the adonis2 function using default settings (999 permutations). Repeated measures correlation analyses were assessed using the *rmcorr package in R (v.0.7.0)*. A two-sided *P* value < 0.05 was considered statistically significant for all analyses performed.

## RESULTS

### Baseline Participants Characteristics

A total of 197 participants (98 women) were included in the present study (**Figure S1**). **Table 1** shows baseline descriptive data of the study participants by group. Fourteen participants (UC: n = 3; early TRE: n = 2; late TRE: n = 4; self-selected TRE: n = 5) were lost during the study due to different reasons (e.g., work conflicts, lack of motivation, or unrelated health/personal issues). However, all 197 participants were included in the intention-to-treat analysis. Sample sizes at baseline, post-intervention, and those included in the linear mixed model analyses are reported for each outcome in **Table S1**. Moreover, liver parameters, adipokines, hepatokines, adipo-hepatokines, and dietary intake at baseline, post-intervention, and their corresponding changes over the 12-week period are presented in **Table S1**.

**Table 1.**
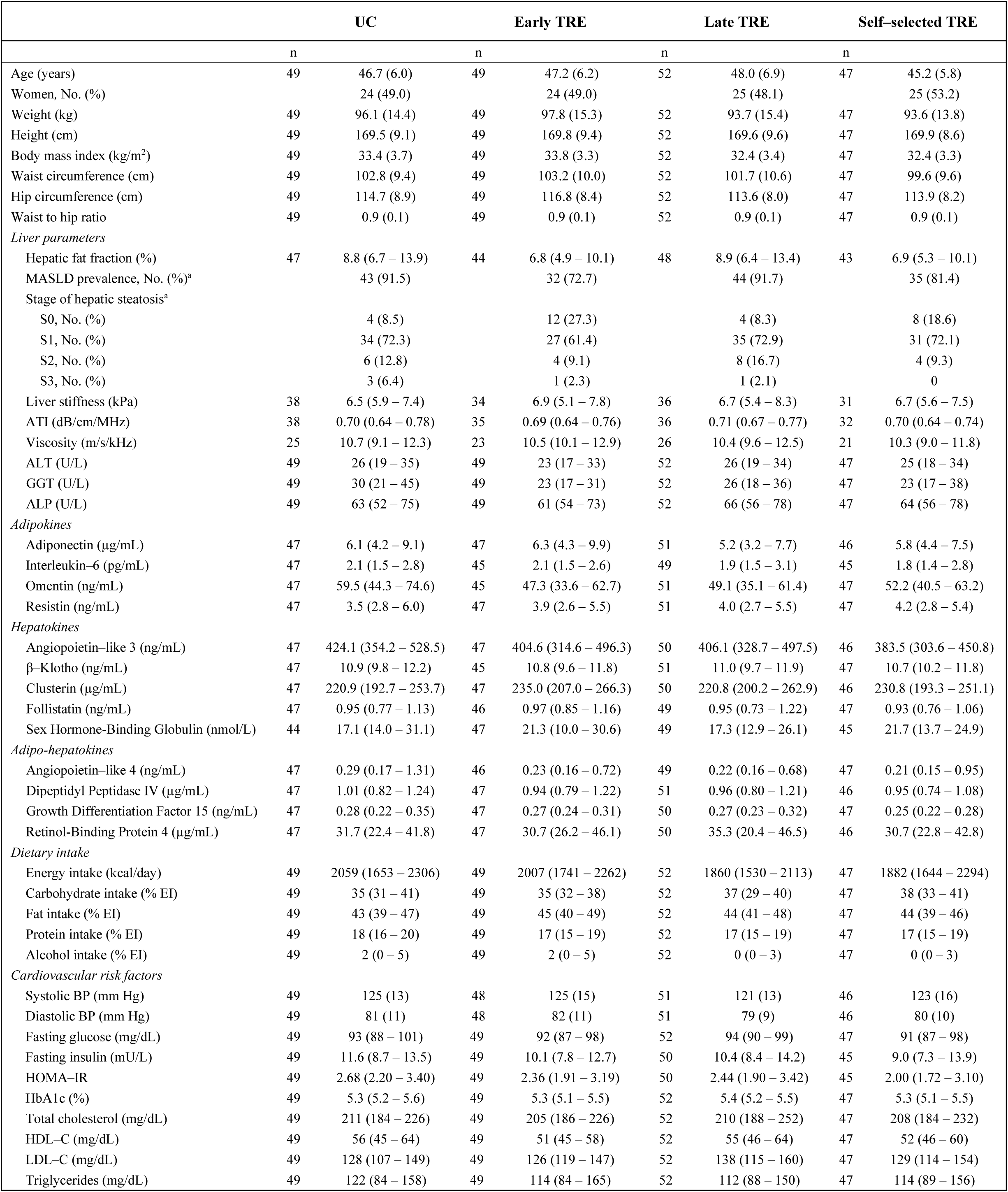
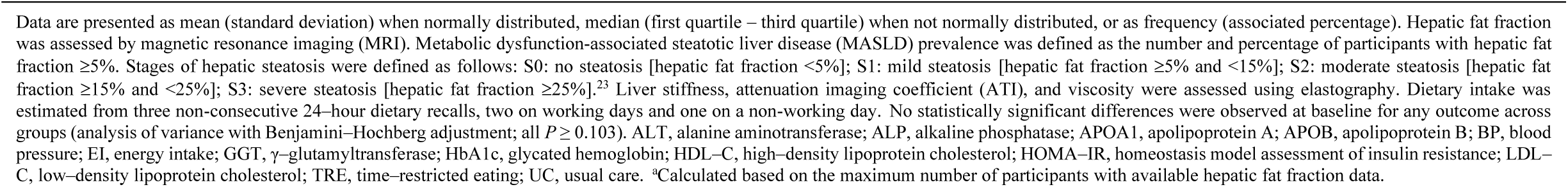
Baseline characteristics of participants by intervention group.

|  | UC |  | Early TRE |  | Late TRE |  | Self-selected TRE |  |
| --- | --- | --- | --- | --- | --- | --- | --- | --- |
|  | n |  | n |  | n |  | n |  |
| Age (years) | 49 | 46.7 (6.0) | 49 | 47.2 (6.2) | 52 | 48.0 (6.9) | 47 | 45.2 (5.8) |
| Women, No. (%) |  | 24 (49.0) |  | 24 (49.0) |  | 25 (48.1) |  | 25 (53.2) |
| Weight (kg) | 49 | 96.1 (14.4) | 49 | 97.8 (15.3) | 52 | 93.7 (15.4) | 47 | 93.6 (13.8) |
| Height (cm) | 49 | 169.5 (9.1) | 49 | 169.8 (9.4) | 52 | 169.6 (9.6) | 47 | 169.9 (8.6) |
| Body mass index (kg/m<sup>2</sup>) | 49 | 33.4 (3.7) | 49 | 33.8 (3.3) | 52 | 32.4 (3.4) | 47 | 32.4 (3.3) |
| Waist circumference (cm) | 49 | 102.8 (9.4) | 49 | 103.2 (10.0) | 52 | 101.7 (10.6) | 47 | 99.6 (9.6) |
| Hip circumference (cm) | 49 | 114.7 (8.9) | 49 | 116.8 (8.4) | 52 | 113.6 (8.0) | 47 | 113.9 (8.2) |
| Waist to hip ratio | 49 | 0.9 (0.1) | 49 | 0.9 (0.1) | 52 | 0.9 (0.1) | 47 | 0.9 (0.1) |
| *Liver parameters* |  |  |  |  |  |  |  |  |
| Hepatic fat fraction (%) | 47 | 8.8 (6.7 – 13.9) | 44 | 6.8 (4.9 – 10.1) | 48 | 8.9 (6.4 – 13.4) | 43 | 6.9 (5.3 – 10.1) |
| MASLD prevalence, No. (%)<sup>a</sup> |  | 43 (91.5) |  | 32 (72.7) |  | 44 (91.7) |  | 35 (81.4) |
| Stage of hepatic steatosis<sup>a</sup> |  |  |  |  |  |  |  |  |
| S0, No. (%) |  | 4 (8.5) |  | 12 (27.3) |  | 4 (8.3) |  | 8 (18.6) |
| S1, No. (%) |  | 34 (72.3) |  | 27 (61.4) |  | 35 (72.9) |  | 31 (72.1) |
| S2, No. (%) |  | 6 (12.8) |  | 4 (9.1) |  | 8 (16.7) |  | 4 (9.3) |
| S3, No. (%) |  | 3 (6.4) |  | 1 (2.3) |  | 1 (2.1) |  | 0 |
| Liver stiffness (kPa) | 38 | 6.5 (5.9 – 7.4) | 34 | 6.9 (5.1 – 7.8) | 36 | 6.7 (5.4 – 8.3) | 31 | 6.7 (5.6 – 7.5) |
| ATI (dB/cm/MHz) | 38 | 0.70 (0.64 – 0.78) | 35 | 0.69 (0.64 – 0.76) | 36 | 0.71 (0.67 – 0.77) | 32 | 0.70 (0.64 – 0.74) |
| Viscosity (m/s/kHz) | 25 | 10.7 (9.1 – 12.3) | 23 | 10.5 (10.1 – 12.9) | 26 | 10.4 (9.6 – 12.5) | 21 | 10.3 (9.0 – 11.8) |
| ALT (U/L) | 49 | 26 (19 – 35) | 49 | 23 (17 – 33) | 52 | 26 (19 – 34) | 47 | 25 (18 – 34) |
| GGT (U/L) | 49 | 30 (21 – 45) | 49 | 23 (17 – 31) | 52 | 26 (18 – 36) | 47 | 23 (17 – 38) |
| ALP (U/L) | 49 | 63 (52 – 75) | 49 | 61 (54 – 73) | 52 | 66 (56 – 78) | 47 | 64 (56 – 78) |
| *Adipokines* |  |  |  |  |  |  |  |  |
| Adiponectin (µg/mL) | 47 | 6.1 (4.2 – 9.1) | 47 | 6.3 (4.3 – 9.9) | 51 | 5.2 (3.2 – 7.7) | 46 | 5.8 (4.4 – 7.5) |
| Interleukin-6 (pg/mL) | 47 | 2.1 (1.5 – 2.8) | 45 | 2.1 (1.5 – 2.6) | 49 | 1.9 (1.5 – 3.1) | 45 | 1.8 (1.4 – 2.8) |
| Omentin (ng/mL) | 47 | 59.5 (44.3 – 74.6) | 45 | 47.3 (33.6 – 62.7) | 51 | 49.1 (35.1 – 61.4) | 47 | 52.2 (40.5 – 63.2) |
| Resistin (ng/mL) | 47 | 3.5 (2.8 – 6.0) | 47 | 3.9 (2.6 – 5.5) | 51 | 4.0 (2.7 – 5.5) | 47 | 4.2 (2.8 – 5.4) |
| *Hepatokines* |  |  |  |  |  |  |  |  |
| Angiopoietin-like 3 (ng/mL) | 47 | 424.1 (354.2 – 528.5) | 47 | 404.6 (314.6 – 496.3) | 50 | 406.1 (328.7 – 497.5) | 46 | 383.5 (303.6 – 450.8) |
| β-Klotho (ng/mL) | 47 | 10.9 (9.8 – 12.2) | 45 | 10.8 (9.6 – 11.8) | 51 | 11.0 (9.7 – 11.9) | 47 | 10.7 (10.2 – 11.8) |
| Clusterin (µg/mL) | 47 | 220.9 (192.7 – 253.7) | 47 | 235.0 (207.0 – 266.3) | 50 | 220.8 (200.2 – 262.9) | 46 | 230.8 (193.3 – 251.1) |
| Follistatin (ng/mL) | 47 | 0.95 (0.77 – 1.13) | 46 | 0.97 (0.85 – 1.16) | 49 | 0.95 (0.73 – 1.22) | 47 | 0.93 (0.76 – 1.06) |
| Sex Hormone-Binding Globulin (nmol/L) | 44 | 17.1 (14.0 – 31.1) | 47 | 21.3 (10.0 – 30.6) | 49 | 17.3 (12.9 – 26.1) | 45 | 21.7 (13.7 – 24.9) |
| *Adipo-hepatokines* |  |  |  |  |  |  |  |  |
| Angiopoietin-like 4 (ng/mL) | 47 | 0.29 (0.17 – 1.31) | 46 | 0.23 (0.16 – 0.72) | 49 | 0.22 (0.16 – 0.68) | 47 | 0.21 (0.15 – 0.95) |
| Dipeptidyl Peptidase IV (µg/mL) | 47 | 1.01 (0.82 – 1.24) | 47 | 0.94 (0.79 – 1.22) | 51 | 0.96 (0.80 – 1.21) | 46 | 0.95 (0.74 – 1.08) |
| Growth Differentiation Factor 15 (ng/mL) | 47 | 0.28 (0.22 – 0.35) | 47 | 0.27 (0.24 – 0.31) | 50 | 0.27 (0.23 – 0.32) | 47 | 0.25 (0.22 – 0.28) |
| Retinol-Binding Protein 4 (µg/mL) | 47 | 31.7 (22.4 – 41.8) | 47 | 30.7 (26.2 – 46.1) | 50 | 35.3 (20.4 – 46.5) | 46 | 30.7 (22.8 – 42.8) |
| *Dietary intake* |  |  |  |  |  |  |  |  |
| Energy intake (kcal/day) | 49 | 2059 (1653 – 2306) | 49 | 2007 (1741 – 2262) | 52 | 1860 (1530 – 2113) | 47 | 1882 (1644 – 2294) |
| Carbohydrate intake (% EI) | 49 | 35 (31 – 41) | 49 | 35 (32 – 38) | 52 | 37 (29 – 40) | 47 | 38 (33 – 41) |
| Fat intake (% EI) | 49 | 43 (39 – 47) | 49 | 45 (40 – 49) | 52 | 44 (41 – 48) | 47 | 44 (39 – 46) |
| Protein intake (% EI) | 49 | 18 (16 – 20) | 49 | 17 (15 – 19) | 52 | 17 (15 – 19) | 47 | 17 (15 – 19) |
| Alcohol intake (% EI) | 49 | 2 (0 – 5) | 49 | 2 (0 – 5) | 52 | 0 (0 – 3) | 47 | 0 (0 – 3) |
| *Cardiovascular risk factors* |  |  |  |  |  |  |  |  |
| Systolic BP (mm Hg) | 49 | 125 (13) | 48 | 125 (15) | 51 | 121 (13) | 46 | 123 (16) |
| Diastolic BP (mm Hg) | 49 | 81 (11) | 48 | 82 (11) | 51 | 79 (9) | 46 | 80 (10) |
| Fasting glucose (mg/dL) | 49 | 93 (88 – 101) | 49 | 92 (87 – 98) | 52 | 94 (90 – 99) | 47 | 91 (87 – 98) |
| Fasting insulin (mU/L) | 49 | 11.6 (8.7 – 13.5) | 49 | 10.1 (7.8 – 12.7) | 50 | 10.4 (8.4 – 14.2) | 45 | 9.0 (7.3 – 13.9) |
| HOMA-IR | 49 | 2.68 (2.20 – 3.40) | 49 | 2.36 (1.91 – 3.19) | 50 | 2.44 (1.90 – 3.42) | 45 | 2.00 (1.72 – 3.10) |
| HbA1c (%) | 49 | 5.3 (5.2 – 5.6) | 49 | 5.3 (5.1 – 5.5) | 52 | 5.4 (5.2 – 5.5) | 47 | 5.3 (5.1 – 5.5) |
| Total cholesterol (mg/dL) | 49 | 211 (184 – 226) | 49 | 205 (186 – 226) | 52 | 210 (188 – 252) | 47 | 208 (184 – 232) |
| HDL-C (mg/dL) | 49 | 56 (45 – 64) | 49 | 51 (45 – 58) | 52 | 55 (46 – 64) | 47 | 52 (46 – 60) |
| LDL-C (mg/dL) | 49 | 128 (107 – 149) | 49 | 126 (119 – 147) | 52 | 138 (115 – 160) | 47 | 129 (114 – 154) |
| Triglycerides (mg/dL) | 49 | 122 (84 – 158) | 49 | 114 (84 – 165) | 52 | 112 (88 – 150) | 47 | 114 (89 – 156) |

### Hepatic fat fraction, liver stiffness, attenuation, viscosity, and liver enzymes

At baseline, MASLD (≥5% hepatic fat fraction) was present in 24 participants (10 women) aged 30–39 years, 77 (34 women) aged 40–49 years, and 53 (22 women) aged 50–60 years, with no significant age-group differences (*P* = 0.22). Overall, men had a higher prevalence of MASLD than women (n = 88 vs. n = 66; *P* < 0.001). By group, MASLD was present in 43 participants (19 women) in the UC group, 32 (10 women) in the early TRE group, 44 (20 women) in the late TRE group, and 35 (17 women) in the self-selected TRE group. Significant reductions in hepatic fat fraction were observed within the early TRE group (mean change: - 1.2%; 95% confidence interval [CI], -2.2 to -0.2; *P* = 0.02), the late TRE group (mean change: -2.3%; 95% CI, -3.3 to -1.3; *P* < 0.001), and the self-selected TRE group (mean change: -1.5%; 95% CI, -2.6 to -0.5; *P* < 0.001), but not in the UC group (mean change: -0.8%; 95% CI, -1.8 to 0.2; *P* = 0.12) (**Table S1**). However, no statistically significant differences were detected in hepatic fat fraction changes between the early TRE (mean difference: -0.4%; 95% confidence interval [CI], -2.3 to 1.5; *P* = 0.95), late TRE (mean difference: -1.5%; 95% CI, -3.3 to 0.3; *P* = 0.15), and self-selected TRE groups (mean difference: -0.7%; 95% CI, -2.6 to 1.2; *P* = 0.77) compared to the UC group (**Figure 1** and **Table 2**). There were no statistically significant differences in hepatic fat fraction changes (i) between the early TRE and late TRE groups (mean difference: 1.1%; 95% CI, -0.7 to 3.0; *P* = 0.41), (ii) between the early TRE and self-selected TRE groups (mean difference: 0.3%; 95% CI, -1.6 to 2.2; *P* = 0.97), and (iii) between the late TRE and self-selected TRE groups (mean difference: -0.8%; 95% CI, -2.7 to 1.1; *P* = 0.69) (**Figure 1** and **Table 3**). The percentage of participants with resolution of MASLD (defined as hepatic fat fraction <5.0%) at 12 weeks was similar across groups (UC: 4.7% [n = 2]; early TRE: 15.6% [n = 5]; late TRE: 6.8% [n = 3]; self-selected TRE: 20.0% [n = 7]; *P* = 0.11). No between-group differences were observed in changes in liver stiffness, attenuation (i.e., ATI), viscosity, or circulating liver enzymes (i.e., ALT, GGT, and ALP) across all groups after the intervention (all *P* ≥ 0.05; **Tables 2 and 3**).

**Figure 1.**
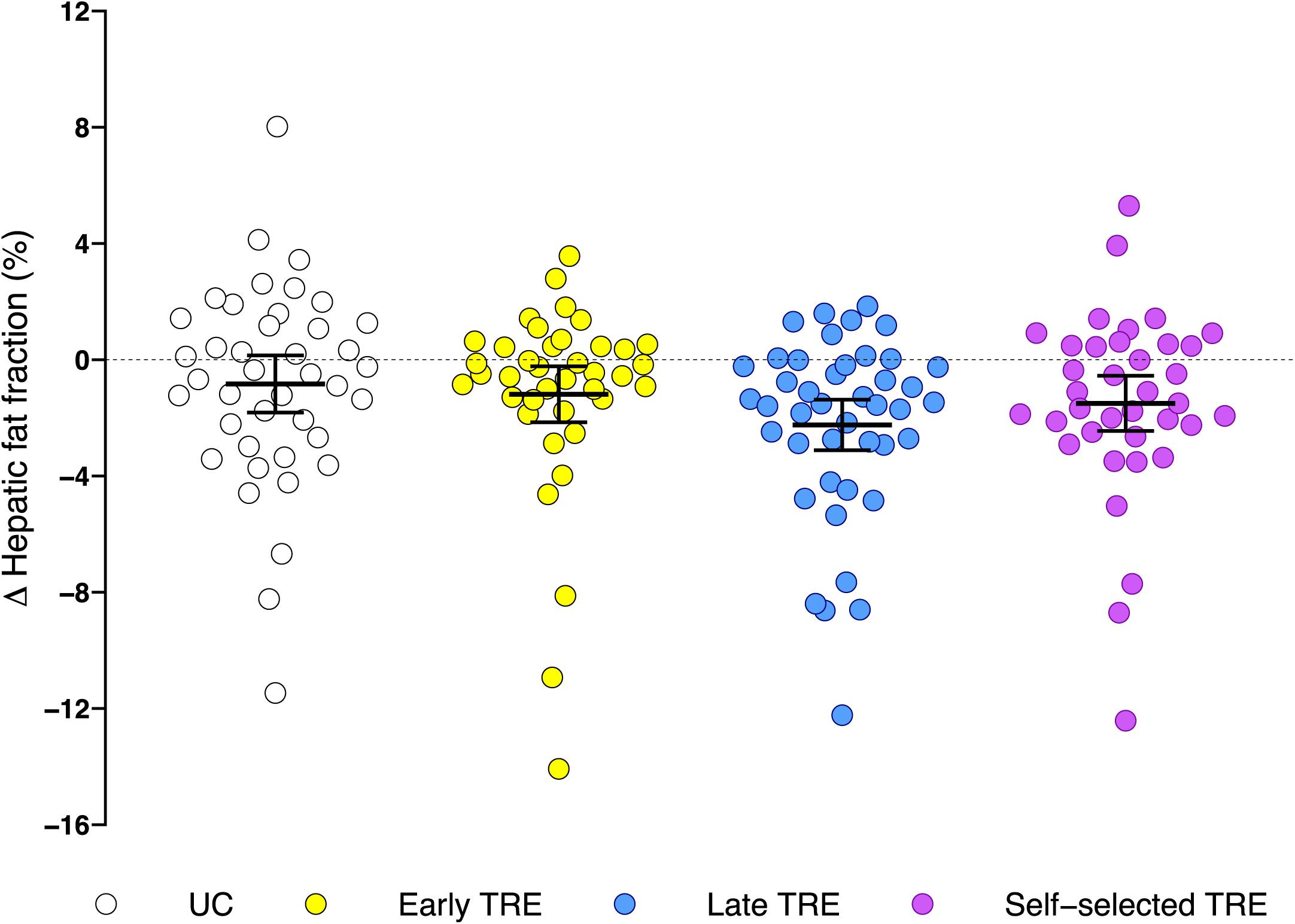
Changes in hepatic fat fraction among the usual care (UC), early time–restricted eating (TRE), late TRE, and self–selected TRE groups after the 12–week intervention. Data are raw means with a 95% confidence interval. No statistically significant differences in changes in hepatic fat fraction were detected across all groups after the intervention. Changes were calculated as postintervention minus preintervention values. Hepatic fat fraction was assessed by magnetic resonance imaging. Intervention effects on hepatic fat fraction at 3 months after the intervention were assessed based on repeated-measures linear mixed-effects multilevel models, which included random cluster (site) effects. Individual measures of change were therefore modelled as a function of randomly assigned group, site, assessment time, and their interaction terms. Sex was also considered a covariate. Model-based estimations were performed with an intention-to-treat approach using the restricted maximum-likelihood method, the model assuming that missing values were missing-at-random.

**Table 2.** Changes in liver parameters, adipokines, hepatokines, adipo-hepatokines, and dietary intake endpoints in the time–restricted eating (TRE) groups compared to the usual care (UC) group after the 12–week intervention.

| Endpoint | Early TRE vs. UC | Late TRE vs. UC | Self-selected TRE vs. UC |
| --- | --- | --- | --- |
|  | Mean difference (95% CI) | Mean difference (95% CI) | Mean difference (95% CI) |
| *Liver parameters* |  |  |  |
| Hepatic fat fraction (%) | -0.4 (-2.3, 1.5) | -1.5 (-3.3, 0.3) | -0.7 (-2.6, 1.2) |
| Liver stiffness (kPa) | 0.4 (-1.1, 1.9) | 0.6 (-0.9, 2.1) | 0.1 (-1.5, 1.6) |
| ATI (dB/cm/MHz) | -0.06 (-0.11, 0.00) | -0.04 (-0.09, 0.02) | -0.03 (-0.09, 0.03) |
| Viscosity (m/s/kHz) | -0.6 (-3.4, 2.1) | 0.5 (-2.1, 3.2) | 0.4 (-2.4, 3.3) |
| ALT (U/L) | -1 (-8, 6) | -3 (-10, 4) | -3 (-10, 4) |
| GGT (U/L) | 1 (-11, 13) | 0 (-11, 12) | 0 (-12, 12) |
| ALP (U/L) | 3 (-3, 8) | -1 (-7, 4) | 0 (-5, 5) |
| *Adipokines* |  |  |  |
| Adiponectin (µg/mL) | -0.5 (-2.9, 1.9) | 0.3 (-2.1, 2.7) | 0.2 (-2.3, 2.7) |
| Interleukin-6 (pg/mL) | -0.3 (-1.3, 0.7) | -0.5 (-1.6, 0.5) | -0.3 (-1.3, 0.8) |
| Omentin (ng/mL) | 0.6 (-14.3, 15.4) | 8.7 (-5.9, 23.2) | 2.5 (-12.4, 17.4) |
| Resistin (ng/mL) | 0.8 (-0.8, 2.3) | 0.0 (-1.5, 1.5) | 0.0 (-1.5, 1.6) |
| *Hepatokines* |  |  |  |
| Angiopoietin-like 3 (ng/mL) | 27.7 (-57.5, 112.9) | 42.9 (-41.6, 127.4) | 42.6 (-43.9, 129.1) |
| β-Klotho (ng/mL) | 0.8 (-1.4, 2.9) | 0.8 (-1.3, 3.0) | 1.6 (-0.6, 3.8) |
| Clusterin (µg/mL) | -1.4 (-43.1, 40.2) | -1.5 (-42.8, 39.7) | -4.3 (-46.6, 37.9) |
| Follistatin (ng/mL) | -0.02 (-0.18, 0.15) | 0.00 (-0.16, 0.16) | 0.01 (-0.15, 0.18) |
| Sex Hormone-Binding Globulin (nmol/L) | 7.1 (-7.0, 21.1) | -0.3 (-14.1, 13.5) | 6.8 (-7.4, 21.0) |
| *Adipo-hepatokines* |  |  |  |
| Angiopoietin-like 4 (ng/mL) | 0.23 (-0.20, 0.67) | 0.27 (-0.16, 0.70) | 0.16 (-0.28, 0.60) |
| Dipeptidyl Peptidase IV (µg/mL) | -0.04 (-0.21, 0.14) | 0.06 (-0.11, 0.23) | 0.05 (-0.12, 0.23) |
| Growth Differentiation Factor 15 (ng/mL) | -0.01 (-0.04, 0.02) | -0.00 (-0.04, 0.03) | 0.01 (-0.02, 0.04) |
| Retinol-Binding Protein 4 (µg/mL) | -1.0 (-9.8, 7.7) | -1.2 (-9.9, 7.5) | 0.0 (-8.9, 8.9) |
| *Dietary intake* |  |  |  |
| Energy intake (kcal/day) | -304 (-565, -44) * | -145 (-404, 115) | -243 (-510, 24) |
| Carbohydrate intake (% EI) | 1 (-3, 5) | 0 (-4, 4) | 0 (-5, 4) |
| Fat intake (% EI) | -1 (-5, 4) | -1 (-6, 3) | -1 (-6, 4) |
| Protein intake (% EI) | 0 (-2, 2) | 0 (-2, 3) | 1 (-1, 3) |
| Alcohol intake (% EI) | 0 (-2, 1) | 1 (-1, 2) | 0 (-1, 2) |

**Table 3.**
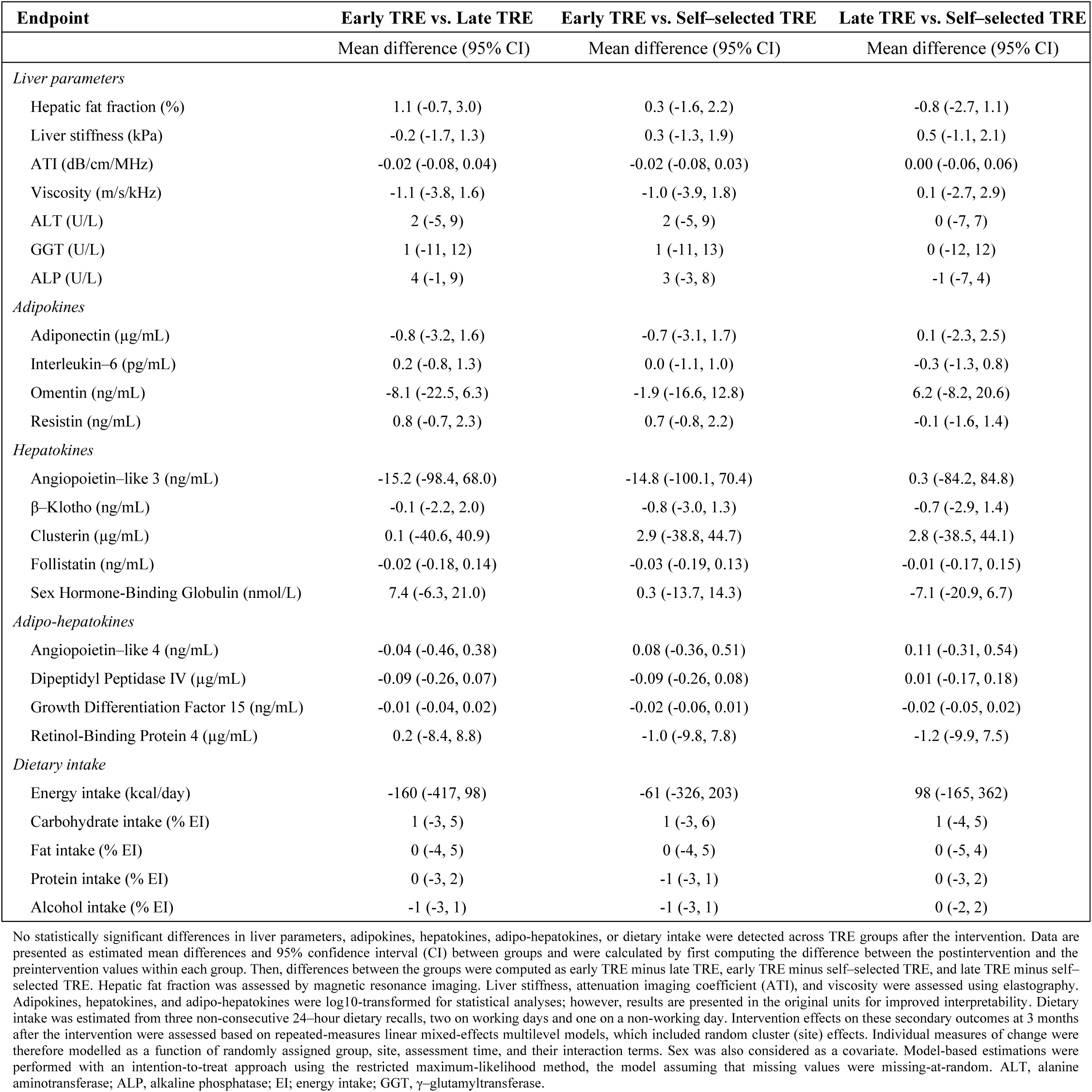
Changes in liver parameters, adipokines, hepatokines, adipo-hepatokines, and dietary intake endpoints in the time–restricted eating (TRE) groups compared to each other after the 12–week intervention.

Interestingly, in exploratory analyses, when participants in the TRE groups were categorized according to presence of MASLD at baseline (≥5% hepatic fat fraction), we observed a significant greater reduction in hepatic fat fraction in those with MALSD compared to those without the condition (mean difference: -2.7%; 95% CI, -4.1 to -1.2; *P* < 0.001; **Figure S6**). These findings remained significant after adjusting for baseline hepatic fat fraction or changes in body weight (both *P* < 0.001).

### Adipokines, hepatokines, adipo-hepatokines, and correlations with changes in hepatic fat fraction

No statistically significant differences in changes in adipokines, hepatokines, and adipo-hepatokines were observed between any TRE group compared with the UC group (all *P* ≥ 0.13; **Figure 2** and **Table 2**), nor among TRE groups (all *P* ≥ 0.24; **Figure 2** and **Table 3**). Repeated measures correlation analyses showed that reductions in hepatic fat fraction over time were significantly associated with decreases in several hepatokines (i.e., ANGPTL3, β–Klotho, clusterin, and follistatin) (all r ≥ 0.263, all *P* = 0.001; **Table S2**), as well as with reductions in selected adipo-hepatokines, specifically DPPIV and RBP4 (both r ≥ 0.307, both *P* < 0.001; **Table S2**) across all sample, and in TRE groups alone (all r ≥ 0.222, all *P* ≤ 0.018; **Table S2**). No additional significant associations were observed between changes in hepatic fat fraction and those obtained in other adipokines, hepatokines, or adipo-hepatokines (all *P* ≥ 0.30; **Table S2**).

**Figure 2.**
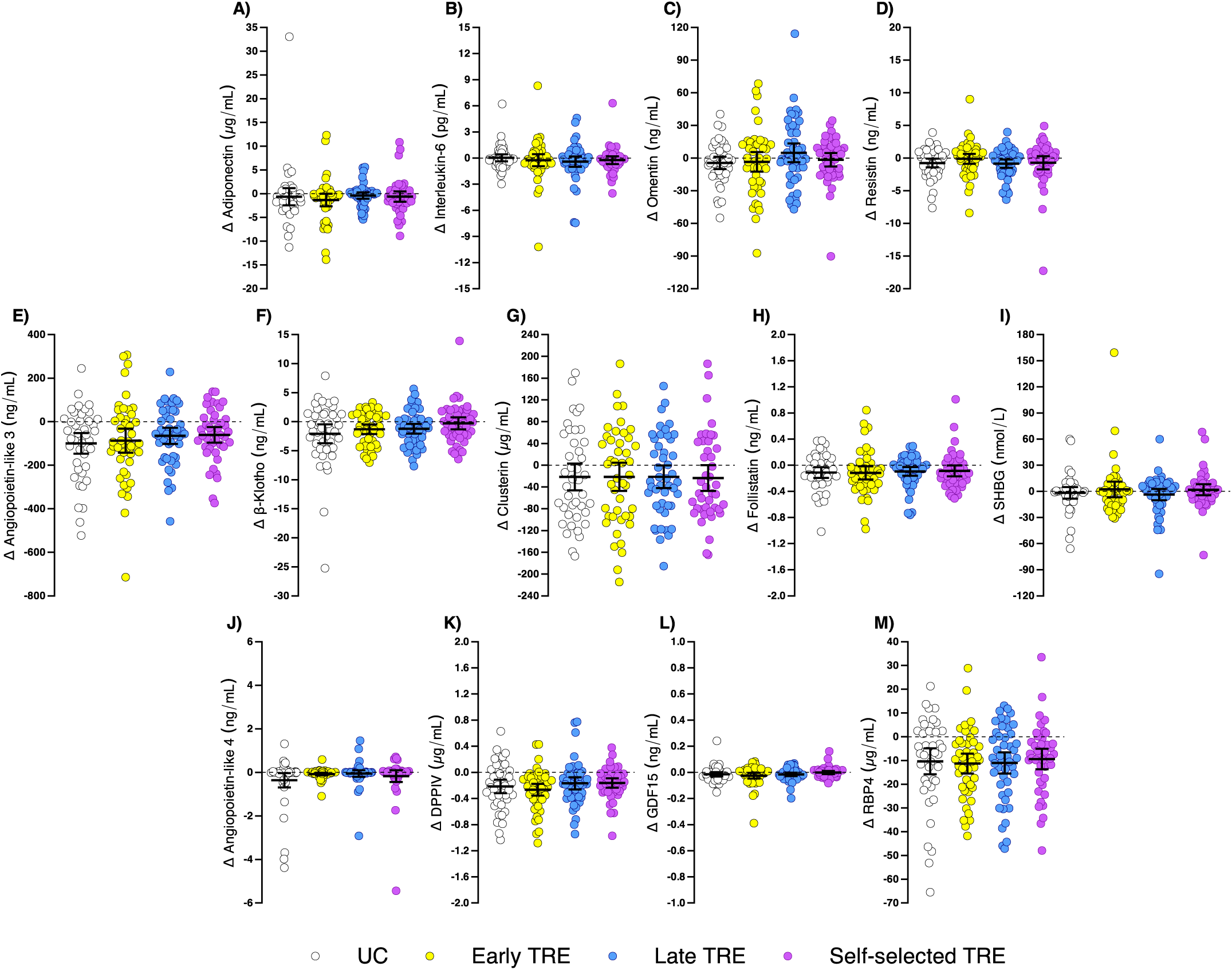
Changes in adiponectin (A), interleukin-6 (B), omentin (C), resistin (D), angiopoietin-like 3 (E), β-Klotho (F), clusterin (G), follistatin (H), sex hormone-binding globulin (SHBG; I), angiopoietin-like 4 (J), dipeptidyl peptidase IV (DPPIV; K), growth differentiation factor 15 (GDF15; L), and retinol-binding protein 4 (RBP4; M) among the usual care (UC), early time–restricted eating (TRE), late TRE, and self–selected TRE groups after the 12–week intervention. Data are raw means with a 95% confidence interval. No statistically significant differences in changes in the distinct adipokines, hepatokines, and adipo-hepatokines were detected across all groups after the intervention. Adipokines, hepatokines, and adipo-hepatokines were log10-transformed for statistical analyses; however, results are presented in the original units for improved interpretability. Changes were calculated as postintervention minus preintervention values. Intervention effects on these secondary outcomes at 3 months after the intervention were assessed based on repeated-measures linear mixed-effects multilevel models, which included random cluster (site) effects. Individual measures of change were therefore modelled as a function of randomly assigned group, site, assessment time, and their interaction terms. Sex was also considered a covariate. Model-based estimations were performed with an intention-to-treat approach using the restricted maximum-likelihood method, the model assuming that missing values were missing-at-random.

### Dietary intake

As reported before,^13^ the decrease in energy intake was significantly greater in the early TRE group (mean difference: -304 kcal/day; 95% CI, -565 to -44; *P* = 0.01) compared with the UC group (**Table 2**). No other significant between-group differences were observed for changes in energy intake, macronutrient intake, or alcohol intake (all *P* ≥ 0.09; **Tables 2 and 3**).

### Correlations between changes in hepatic fat fraction and changes in insulin resistance

As reported before,^13^ no statistically significant differences were detected in changes in insulin resistance, as assessed by HOMA-IR, across all groups after the intervention (all *P* ≥ 0.86). However, we observed that reductions in hepatic fat fraction after the intervention were significantly associated with decreases in HOMA-IR across all sample (r = 0.416; *P* < 0.001; **Table S2**), and in TRE groups alone (r = 0.360, *P* < 0.001; **Table S2**).

### Fecal microbiota composition

Alpha and beta diversity indices of fecal microbiota before and after the intervention are shown in **Table S3** and **Figure S3A-D**, respectively. No statistically significant differences were observed neither in baseline fecal microbiota composition (*P* = 0.29), or beta (*P* = 0.30) and alpha (all *P* ≥ 0.42) diversity indices across all groups (**Figure S2**). There were no statistically significant differences between groups in changes in alpha and beta diversity indices during the intervention (all *P* ≥ 0.16; **Figure 3A-E**). The UC group showed the highest number of taxa that changed during the 3-month intervention (**Figure S4A**). No statistically significant differences were identified when specific changes in fecal microbiota composition between groups were evaluated through MaAsLin3 analysis after FDR adjustment (all *P* ≥ 0.60; **Figure 3F-G and Figure S3E**). **Figure S5** shows within-group LEfSe analyses for metabolic pathway biomarker discovery. MaAsLin3 analysis revealed no significant between-group differences in changes in bacterial functionality during the intervention after FDR adjustment (all *P* ≥ 0.99).

**Figure 3.**
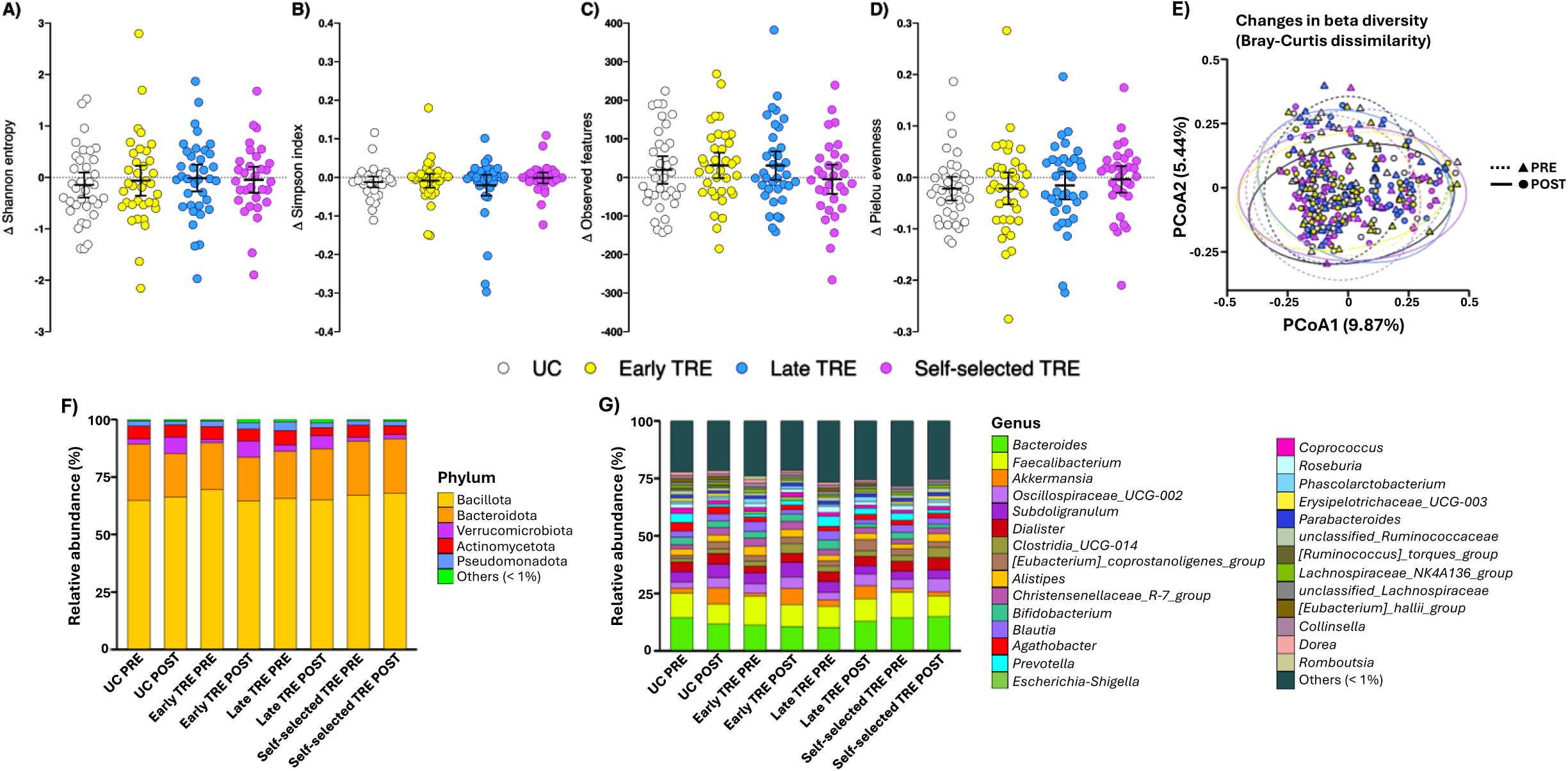
Changes in diversity and microbial taxonomic composition among the usual care (UC), early time–restricted eating (TRE), late TRE, and self–selected TRE groups after the 12–week intervention. **(A-E)** Comparisons between groups for changes in alpha and beta diversity indices during the intervention: **(A)** Shannon entropy (all group comparisons *P* ≥ 0.88), **(B)** Simpson index (all group comparisons *P* ≥ 0.50), **(C)** observed features (all group comparisons *P* ≥ 0.49), **(D)** Pielou evenness (all group comparisons *P* ≥ 0.78), and **(E)** Bray-Curtis dissimilarity (all group comparisons *P* ≥ 0.16). No statistically significant differences were detected in changes in alpha and beta diversity indices across all groups after the intervention. Changes in alpha diversity **(A-D)** were calculated as the difference between the postintervention and the preintervention values. Intervention effects on these secondary outcomes at 3 months after the intervention were assessed based on repeated-measures linear mixed-effects multilevel models, which included random cluster (site) effects. Individual measures of change were therefore modelled as a function of randomly assigned group, site, assessment time, and their interaction terms. Sex was also considered as a covariate. Changes in beta diversity **(E)** were assessed using PERMANOVA, considering the assigned group, assessment time, and their interactions terms. **(F-G)** Microbial taxonomic composition at the phylum **(F)** and genus level **(G)** in fecal samples for the preintervention and postintervention time points for each intervention group. The bars show the mean proportion of relative abundance. The “Others” category in the graphs represents the aggrupation of bacterial groups with less than 1% relative abundance. Changes in bacterial composition were evaluated through MaAsLin3 algorithm considering the assigned group, assessment time, and their interactions terms with sex as a covariate. No statistically significant differences were detected after applying Benjamini-Hochberg False Discovery Rate (FDR). Statistical significance was set at *P* < 0.05. PRE, preintervention; POST, postintervention.

Changes in hepatic fat fraction were not associated with changes in fecal microbiota diversity metrics or composition (all *P* ≥ 0.22). In addition, when TRE participants were grouped according to the baseline presence of MASLD, no differences were found in changes in fecal microbiota diversity or composition throughout the intervention (all *P* ≥ 0.65).

### Eating window times and adherence

As reported before,^13^ the eating window was significantly shorter in the early TRE group (median: 7.7 h; interquartile range, 0.5; *P* < 0.001), the late TRE group (median: 7.4 h; interquartile range, 0.8; *P* < 0.001), and the self-selected TRE group (median: 7.6 h; interquartile range, 0.4; *P* < 0.001) compared to the UC group (median: 13.4 h; interquartile range, 1.2). Median eating window times were 08:30 to 22:00 for the UC group, 09:45 to 17:30 for the early TRE group, 14:20 to 21:30 for the late TRE group, and 12:20 to 20:00 for the self-selected TRE group. The percentage of adherent days (defined as an eating window ≤9 h) was comparable across the early TRE group (mean: 85.3%; 95% CI, 80.6 to 90.0), late TRE group (mean: 88.1%; 95% CI, 84.9 to 91.3) and self-selected TRE group (mean: 85.4%; 95% CI, 81.3 to 89.5) (all *P* ≥ 0.99).

### Associations of clinically meaningful weight loss with changes in hepatic fat fraction, liver stiffness, attenuation, viscosity, liver enzymes, adipokines, hepatokines, adipo-hepatokines, and dietary intake

Descriptive characteristics of participants who lost <5% or ≥5% of their initial body weight are presented in **Table S4**. The proportion of participants who achieved clinically meaningful body weight loss (≥5%; so-called responders) significantly differed among the groups (UC = 16%, early TRE = 41%, late TRE = 44%, self-selected = 43%; *P* = 0.001; **Figure S7**). Specifically, those who achieved a reduction in body weight of at least 5% showed a greater decrease in hepatic fat fraction (mean difference: -2.6%; 95% CI, -3.5 to -1.6; *P* < 0.001; **Figure S8 and Table S5**) and ALT (mean difference: -9 mg/dL; 95% CI, -13 to -5; *P* < 0.001; **Table S5**), yet no other significant differences were observed in changes in liver stiffness, ATI, or other liver enzyme levels (all *P* ≥ 0.21; **Table S5**). The percentage of participants with resolution of MASLD at 12 weeks was similar in both groups (<5% body weight loss: 7.5% [n = 7]; ≥5% body weight loss: 18.9% [n = 10]; *P* = 0.09). Moreover, participants losing ≥5% of their initial body weight had a statistically significant reduction on the adipokine omentin (mean difference: -9.8 ng/mL; 95% CI, -17.8 to -1.7; *P* = 0.046; **Figure S9C and Table S5**), and the hepatokines ANGPTL3 (mean difference: -48.8 ng/mL; 95% CI, -96.1 to -1.5; *P* = 0.046; **Figure S9E and Table S5**) and clusterin (mean difference: -32.5 µg/mL; 95% CI, -55.4 to - 9.6; *P* = 0.02; **Figure S9G and Table S5**). No additional significant differences were noted in adipokines, hepatokines, and adipo-hepatokines changes between participants who lost ≥5% compared to those who lost <5% (all *P* ≥ 0.35; **Figure S9 and Table S5**). No statistically significant between-group differences were observed in changes in energy intake, macronutrient intake, or alcohol intake (all *P* ≥ 0.13**; Table S5**).

### Sensitivity analyses: effects of the intervention by sex

Of note is that we repeated all the analyses separately in men and women, and the results consistently showed similar effects across the TRE groups compared with UC and among the TRE groups themselves (data not shown).

## DISCUSSION

This secondary analysis of RCT suggests that adding TRE, regardless of the timing of the eating window, to a Mediterranean diet-based nutritional education program (UC) for 12 weeks did not offer any additional benefits over UC alone for reducing hepatic fat fraction, improving liver health markers, or modulating fecal microbiota composition in adults with overweight or obesity. It is worth noting that participants from TRE groups with MASLD at baseline showed larger reductions in hepatic fat fraction than those without the condition, which may indicate that TRE could be more effective in individuals with pre-existing hepatic steatosis, consistent with recent studies.^38^ In exploratory analyses, we observed that participants who achieved clinically significant weight loss (≥5%), whom 91% were from the TRE groups, exhibited greater reductions in hepatic fat fraction, along with improvements in ALT, the adipokine omentin, and the hepatokines ANGPTL3 and clusterin, compared to those who did not. These findings raise the possibility that improvements in hepatic fat fraction and liver health markers may be more closely related to the extent of weight loss than by the eating window timing.

Our findings are broadly consistent with previous trials investigating the effects of TRE on hepatic fat fraction measured by MRI. Deng et al.^15^ reported similar reductions in hepatic fat fraction and improvements in liver health markers when comparing 8-hour early TRE (eating window from 08:00 to 16:00) versus late TRE (12:00 to 20:00) after 8 weeks in patients with overweight and MASLD. In contrast, Andriessen et al.^17^ showed no significant changes in hepatic fat fraction after a 3-week TRE intervention with a 10-hour eating window ending before 18:00 in individuals with type 2 diabetes, suggesting that longer intervention durations may be required. Wei et al.^16^ conducted a 12-month trial in patients with obesity and MASLD concluding that early TRE combined with calorie restriction led to comparable improvements in hepatic fat fraction and liver health markers compared with calorie restriction alone. Similarly, the CHRONO-NAFLD Project found no differences in liver steatosis or stiffness (assessed by liver elastography) between 10-hour eating window early TRE, late TRE, and an unrestricted eating window, all combined with a hypocaloric Mediterranean diet, in patients with MASLD over 12 weeks.^39^ These studies align closely with our findings in adults with overweight or obesity, supporting the idea that energy deficit—reflected in the greater reduction in energy intake observed in the early TRE group compared with the UC group—rather than eating window timing, could be the main determinant for improvements in liver-related outcomes. Nevertheless, we observed slightly higher MASLD remission rates in the early and self-selected TRE groups compared to the UC and late TRE groups, although these differences did not reach statistical significance. Potential explanations could include the alignment of early or self-selected eating windows with circadian rhythms, which may facilitate improved metabolic regulation; indeed, we previously reported in this cohort that early TRE reduced fasting and nocturnal glucose levels compared with UC, late TRE, and self-selected TRE groups.^13^ This pattern suggests that late TRE may be less effective in reducing hepatic fat fraction, a possibility that warrants further investigation. A particularly noteworthy finding was that participants in the TRE groups with pre-existing MASLD at baseline experienced a significantly greater reduction in hepatic fat fraction than those without the condition (-2.1% vs. +0.5%). This finding may support the notion that TRE can be especially effective in individuals with established hepatic steatosis. This observation aligns with recent studies ^38^ showing that individuals with elevated baseline hepatic fat levels exhibit a more pronounced response to TRE, possibly due to a greater capacity for metabolic and hepatic improvement. Moreover, the observed associations of reductions in hepatic fat fraction with decreases in insulin resistance, HOMA-IR, reinforce the interconnection between hepatic steatosis and metabolic dysfunction in the progression of MASLD.^40^ Consequently, these findings underscore the potential of TRE as a targeted and promising intervention for managing early-stage MASLD.

A higher proportion of participants in the TRE groups achieved clinically meaningful body weight loss (≥5%) compared to the UC group. Our exploratory analysis showed that those who lost ≥5% of their initial body weight experienced significantly greater reductions in hepatic fat fraction (-3.1% vs. -0.5%) and ALT, as well as improvements in several circulating biomarkers, including the hepatokines ANGPTL3 and clusterin, and the adipokine omentin, compared to those who did not. These changes suggest a beneficial modulation of hepatokine secretion patterns associated with the resolution of hepatic steatosis.^41^ Although MASLD remission rates did not differ between responders and non-responders, achieving ≥5% weight loss was associated with a modest reduction in hepatic fat fraction (−3.1%), consistent with recent evidence suggesting that at least 5% body weight loss can produce clinically meaningful reductions in liver steatosis.^40^ Given the emerging understanding that fatty liver alters its endocrine function by increasing hepatokines such as ANGPTL3 and clusterin,^41^ our results suggest that body weight loss may lead to a healthier hepatokine profile, thereby improving systemic metabolic health. However, these effects were not evident when comparing the UC group to the TRE groups, highlighting the importance of the degree of weight loss in driving liver health benefits. This hypothesis is further supported by the temporal relationship observed between reductions in hepatic fat fraction and declines in ANGPTL3, β-Klotho, clusterin, DPPIV, and RBP4. In this context, TRE may primarily act as an effective strategy to facilitate body weight loss, with the achieved energy deficit being the key mediator of liver health improvements rather than the specific timing of the eating window in individuals with overweight or obesity. Nonetheless, other pathophysiological mechanisms may also contribute to the observed improvements. These could include reductions in subclinical inflammation, enhanced hepatic insulin sensitivity, alterations in lipid metabolism, and modulation of the gut–liver axis beyond changes in microbiota composition. Future trials with mechanistic endpoints will be important to disentangle the relative contributions of these pathways from the effects of weight loss alone.

One of the proposed mechanisms through which TRE could exert metabolic benefits, specifically in improving obesity and MASLD, is through the modulation of the gut microbiota, which is associated with the established gut-liver axis linking gut and liver health.^42^ However, the exact impact of TRE on the microbiome remains poorly understood, with varying results reported in previous studies.^43–48^ In our study, we did not find significant changes in fecal microbiota composition or in alpha and beta diversity due to TRE when compared to the UC group. Similarly, we observed no association between changes in hepatic fat fraction and alterations in fecal microbiota, which may be attributable to the limited impact of TRE on the gut microbiota. Our findings are in accordance with other studies investigating TRE interventions in adults with obesity. Johnson et al.^43^ and Gabel et al.^44^ reported no significant changes in gut microbiome composition and diversity after a 12-week self-selected 8-hour TRE intervention (n = 16 and n = 14 participants, respectively). Similarly, Ferrocino et al.^45^ reported similar findings after a 12-week intervention with an eating window of <12 hours, compared to a control group (n = 25 and n = 24, respectively), with both groups undergoing similar caloric restriction. Our study reinforces these findings by addressing some of the limitations of previous research, such as a larger sample size, a controlled study design, a sex-balanced sample, and the comparison of different TRE schedules. Further studies are still needed to understand the effect of different TRE schedules on gut microbiota, considering factors such as the timing of sample collection,^49^ the time of the last meal, and the method of analysis (e.g., 16S amplicon sequencing versus shotgun metagenomics).

### Limitations of the study

This study presents several limitations. It is a secondary analysis, and therefore the power calculation of sample size was not based on hepatic fat fraction, the other health-related liver health markers, or fecal microbiota composition, but on visceral adipose tissue changes,^13^ the main outcome of the study. It should also be noted that participants had normal or only mildly elevated hepatic fat fraction, liver stiffness, liver enzymes, and cardiometabolic risk factors at baseline, which may have reduced the potential to detect larger effects compared with trials that enrolled participants with MASLD or more advanced metabolic disease. This is highlighted by our analysis showing that participants with MASLD at baseline experienced greater improvements in hepatic fat fraction than those without the condition. Additionally, the 12-week intervention period may not be long enough to observe significant differences between intervention groups, as well as to induce changes in fecal microbiota. Furthermore, although the methodology used for microbiota analyses was robust, reliance on 16S rRNA profiling rather than shotgun metagenomics may have limited the detection of functional microbial shifts. The absence of a non-intervention control group also restricts interpretation of hepatic fat, liver health, and fecal microbiota outcomes. Finally, emerging evidence suggests that fecal microbiota composition is influenced by circadian rhythms and that the timing of stool collection may affect replicability.^49^ Because stool samples in our study were not collected at standardized times, this could represent a potential limitation. In addition, the findings may not be generalizable beyond the Spanish population due to cultural differences in meal timing, as Spain is characterized by a later meal schedule compared with other countries.

### Conclusions

In this secondary analysis of a RCT, which implemented three different TRE schedules, the findings suggest that adding TRE to a Mediterranean diet-based nutritional program does not confer additional benefits for reducing hepatic fat fraction, improving liver health markers or fecal microbiota composition beyond those achieved with dietary education alone in men and women with overweight or obesity. Our exploratory analysis suggests that TRE is a highly promising and targeted strategy for managing early stages of MASLD, given its demonstrated ability to induce significant reductions in hepatic fat fraction in individuals with pre-existing hepatic steatosis. Of note is that participants who achieved a clinically meaningful weight loss (≥5%)—the majority of whom belonged to the TRE groups (91%)—exhibited greater reductions in hepatic fat fraction, along with improvements in ALT, omentin, ANGPTL3, and clusterin levels, compared to those who did not. These preliminary findings suggest that liver-related improvements may be more closely linked to the degree of weight loss than to eating window timing, with TRE potentially contributing by facilitating weight reduction. These results should, however, be interpreted with caution, as the study was not powered for hepatic or microbiota outcomes. Further adequately powered trials are warranted to corroborate our results and determine whether the timing of the eating window, particularly early versus late TRE, differentially impacts hepatic fat fraction, liver health markers, and underlying mechanisms, with a focus on the gut–liver axis. Moreover, further research is needed in populations with MASLD, cirrhosis, and those with chronic diseases, where circadian factors may play a key role.

## Data Availability

Due to privacy concerns, the datasets used in this study are not publicly available. However, researchers can request access to specific individual-level data for academic use only, within 36 months following the publication date, after de-identification. Proposals should be directed to. Upon proposal acceptance, requestors will be granted access to the data after signing a data access agreement. In addition, raw 16S rRNA gene amplicon sequencing data are available from NCBI Sequence Read Archive (SRA) repository under BioProject PRJNA1287213, access at:
https://dataview.ncbi.nlm.nih.gov/object/PRJNA1287213?reviewer=v3a70vcokbh99m4nbrejbjk5h8

https://dataview.ncbi.nlm.nih.gov/object/PRJNA1287213?reviewer=v3a70vcokbh99m4nbrejbjk5h8

## ACKNOWLEDGMENTS

The authors thank all the participants who took part in the study. This study (Project ref. PID2022.141506OB.I00) received support by the MCIU/AEI /10.13039/501100011033 and by ERDF, EU a way of making Europe to Dr Ruiz; The Junta de Andalucía, Consejería de Transformación económica, Industria, Conocimiento y Universidades (A-CTS-516-UGR20) to Dr Ruiz; the University of Granada Plan Propio de Investigación-Excellence actions: Unit of Excellence on Exercise Nutrition and Health (UCEENS) to Dr Ruiz; Spanish Ministry of Universities (FPU18/03357 to Dr Dote-Montero and FPU21/01161 to Mr Clavero-Jimeno) – University of Granada; the Government of Navarra, Departamento de Desarrollo Economico y Empresarial (0011-1365-2021-00070), Plan de Promoción de Grupos de Investigación de la Universidad Pública de Navarra to Dr Labayen, Juan de la Cierva Formación (FJC2020-044453-I to Dr Camacho-Cardenosa) funded by the Ministerio de Ciencia e Innovación and by the European Union NextGenerationEU/PRTR. Dr Alfaro-Magallanes is supported by a Margarita Salas grant RCMS-22-KIAO1C-24-GSMTDX, which is funded by the MCIN/AEI/10.13039/501100011033 and the European Union. Dr Cortés-Martín is supported by JDC2022-048679-I, funded by MCIN/AEI/10.13039/501100011033 and by the European Union “NextGenerationEU”/PRTR. Dr Osés is supported by the Spanish Ministry of Economy and Competitiveness (BES-2017-080770). In addition, funding was provided from the EXERNET Research Network on Exercise and Health (DEP2005-00046/ACTI; 09/UPB/19; 45/UPB/20; 27/UPB/21 to Drs Labayen and Ruiz). This work is part of a doctorate thesis conducted in the Official Doctoral Program in Biomedicine of the University of Granada, Spain.

## AUTHOR CONTRIBUTIONS

Conceptualization: M.D.M., I.L., J.R.R.

Methodology: M.D.M., I.L., J.R.R.

Project administration: M.D.M., A.C.J., E.M.R., M.O., A.C.C., I.L., J.R.R.

Investigation: M.D.M., A.C.J., A.C.M., A.L.P., E.M.R., A.C.C., M.C., M.O., A.L.V.,

F.J.A.G., P.V.G.P., J.G., A.R.N., F.G., C.M.H., M.J.V.

Formal analysis: M.D.M., A.C.J., A.C.M., J.R.R.

Resources: M.A.A., J.G., A.R.N., F.G., J.L.M.R., R.C., M.M.T., I.L., J.R.R.

Data curation: M.D.M., A.C.J., A.C.M., A.L.P., E.M.R., A.C.C., M.C., M.O., M.A.A., A.L.V., J.G., A.R.N., F.G., C.M.H., M.J.V., V.M.A.M., R.C.

Writing – Original Draft: M.D.M., A.C.J., A.C.M., J.R.R.

Writing – Review & Editing: All authors

Supervision: I.L., M.M.T., J.R.R.

Funding acquisition: I.L., M.M.T., J.R.R.

All authors had access to the study data and reviewed and approved the final manuscript.

## DATA TRANSPARENCY STATEMENT

Due to privacy concerns, the datasets used in this study are not publicly available. However, researchers can request access to specific individual-level data for academic use only, within 36 months following the publication date, after de-identification. Proposals should be directed to. Upon proposal acceptance, requestors will be granted access to the data after signing a data access agreement. In addition, raw 16S rRNA gene amplicon sequencing data are available from NCBI Sequence Read Archive (SRA) repository under BioProject PRJNA1287213, access at: https://dataview.ncbi.nlm.nih.gov/object/PRJNA1287213?reviewer=v3a70vcokbh99m4nbrejbjk5h8

