## Supplementary material for "Early, Late, & Self-Selected Time-Restricted Eating: Impact on Hepatic Fat, Liver Health, & Fecal Microbiota in Adults with Overweight or Obesity": Figure S1

*SUPPLEMENTAL INFORMATION*

**Table S1**. Liver parameters, adipokines, hepatokines, adipo-hepatokines, and dietary intake endpoints at baseline and after the 12–week intervention in each intervention group.

| **Endpoint** | **UC** | | **Early TRE** | | **Late TRE** | | **Self-selected TRE** | |
|---|---|---|---|---|---|---|---|---|
| | n | Mean (95% CI) | n | Mean (95% CI) | n | Mean (95% CI) | n | Mean (95% CI) |
| *Liver parameters* | | | | | | | | |
| Hepatic fat fraction (%) | | | | | | | | |
| Preintervention | 47 | 10.9 (9.4, 12.3) | 44 | 8.8 (7.4, 10.3) | 48 | 10.7 (9.3, 12.1) | 43 | 8.5 (7.0, 9.9) |
| Postintervention | 42 | 10.1 (8.6, 11.5) | 41 | 7.7 (6.2, 9.1) | 46 | 8.4 (7.0, 9.8) | 39 | 7.0 (5.4, 8.5) |
| Change | 48 | -0.8 (-1.8, 0.2) | 46 | -1.2 (-2.2, -0.2) * | 51 | -2.3 (-3.3, -1.3) * | 45 | -1.5 (-2.6, -0.5) * |
| Liver stiffness (kPa) | | | | | | | | |
| Preintervention | 38 | 7.1 (6.3, 7.9) | 34 | 7.1 (6.3, 7.9) | 36 | 7.5 (6.7, 8.3) | 31 | 6.9 (6.0, 7.7) |
| Postintervention | 35 | 6.7 (5.9, 7.5) | 37 | 7.1 (6.3, 7.9) | 40 | 7.6 (6.9, 8.4) | 35 | 6.6 (5.7, 7.4) |
| Change | 43 | -0.4 (-1.2, 0.4) | 42 | 0.0 (-0.8, 0.8) | 46 | 0.2 (-0.6, 1.0) | 43 | -0.3 (-1.2, 0.6) |
| ATI (dB/cm/MHz) | | | | | | | | |
| Preintervention | 38 | 0.71 (0.68, 0.75) | 35 | 0.72 (0.69, 0.76) | 36 | 0.74 (0.71, 0.77) | 32 | 0.70 (0.66, 0.73) |
| Postintervention | 35 | 0.70 (0.67, 0.73) | 37 | 0.66 (0.62, 0.69) | 40 | 0.69 (0.66, 0.72) | 35 | 0.65 (0.62, 0.68) |
| Change | 43 | -0.01 (-0.04, 0.02) | 43 | -0.07 (-0.10, -0.04) * | 46 | -0.05 (-0.08, -0.02) * | 43 | -0.04 (-0.08, -0.01) * |
| Viscosity (m/s/kHz) | | | | | | | | |
| Preintervention | 25 | 11.1 (9.9, 12.2) | 23 | 11.8 (10.6, 13.0) | 26 | 11.1 (10.0, 12.2) | 21 | 10.8 (9.6, 12.1) |
| Postintervention | 24 | 11.0 (9.8, 12.2) | 23 | 11.1 (9.9, 12.3) | 26 | 11.5 (10.4, 12.7) | 20 | 11.1 (9.9, 12.4) |
| Change | 25 | -0.1 (-1.5, 1.4) | 23 | -0.7 (-2.2, 0.8) | 27 | 0.4 (-1.0, 1.8) | 22 | 0.3 (-1.2, 1.9) |
| ALT (U/L) | | | | | | | | |
| Preintervention | 49 | 30 (25, 34) | 49 | 28 (24, 33) | 52 | 28 (24, 33) | 47 | 27 (23, 32) |
| Postintervention | 44 | 28 (24, 33) | 47 | 25 (21, 30) | 48 | 24 (19, 28) | 43 | 22 (18, 27) |
| Change | 49 | -2 (-6, 2) | 49 | -3 (-7, 1) | 52 | -5 (-9, -1) * | 47 | -5 (-9, -1) * |
| GGT (U/L) | | | | | | | | |
| Preintervention | 49 | 48 (35, 61) | 49 | 28 (15, 41) | 52 | 35 (22, 47) | 47 | 29 (16, 43) |
| Postintervention | 44 | 44 (31, 57) | 47 | 25 (12, 38) | 48 | 31 (18, 43) | 43 | 25 (12, 38) |
| Change | 49 | -5 (-11, 2) | 49 | -3 (-10, 3) | 52 | -4 (-10, 2) | 47 | -4 (-11, 2) |
| ALP (U/L) | | | | | | | | |
| Preintervention | 49 | 66 (61, 70) | 49 | 65 (61, 70) | 52 | 68 (64, 73) | 47 | 66 (61, 71) |
| Postintervention | 44 | 66 (61, 71) | 47 | 68 (63, 73) | 48 | 67 (63, 72) | 43 | 66 (61, 71) |
| Change | 49 | 0 (-2, 3) | 49 | 3 (0, 6) * | 52 | -1 (-4, 2) | 47 | 0 (-3, 3) |
| *Adipokines* | | | | | | | | |
| Adiponectin (μg/mL) | | | | | | | | |
| Preintervention | 47 | 7.6 (6.1, 9.0) | 47 | 7.8 (6.4, 9.3) | 51 | 5.7 (4.3, 7.1) | 46 | 7.0 (5.5, 8.5) |
| Postintervention | 43 | 6.8 (5.4, 8.3) | 47 | 6.6 (5.2, 8.1) | 48 | 5.3 (3.9, 6.7) | 43 | 6.5 (5.0, 8.0) |
| Change | 49 | -0.7 (-2.0, 0.6) * | 49 | -1.2 (-2.5, 0.1) | 52 | -0.4 (-1.7, 0.9) | 47 | -0.5 (-1.8, 0.8) |
| Interleukin–6 (pg/mL) | | | | | | | | |
| Preintervention | 47 | 2.2 (1.8, 2.7) | 45 | 2.5 (2.1, 3.0) | 49 | 2.7 (2.2, 3.1) | 45 | 2.2 (1.7, 2.7) |
| Postintervention | 44 | 2.3 (1.8, 2.7) | 47 | 2.3 (1.8, 2.7) | 48 | 2.2 (1.7, 2.6) | 43 | 2.0 (1.5, 2.4) |
| Change | 48 | 0.0 (-0.5, 0.6) | 48 | -0.3 (-0.8, 0.3) | 50 | -0.5 (-1.0, 0.0) * | 47 | -0.2 (-0.8, 0.3) |
| Omentin (ng/mL) | | | | | | | | |
| Preintervention | 47 | 79.5 (63.4, 95.6) | 45 | 56.4 (40.2, 72.5) | 51 | 50.3 (34.7, 65.8) | 47 | 55.2 (38.8, 71.5) |
| Postintervention | 43 | 75.6 (59.4, 91.9) | 47 | 53.1 (37.0, 69.1) | 48 | 55.0 (39.4, 70.7) | 43 | 53.8 (37.3, 70.3) |
| Change | 49 | -3.9 (-12.0, 4.2) | 49 | -3.3 (-11.2, 4.6) | 52 | 4.8 (-2.8, 12.4) | 47 | -1.4 (-9.3, 6.5) |
| Resistin (ng/mL) | | | | | | | | |
| Preintervention | 47 | 4.6 (4.0, 5.3) | 47 | 4.4 (3.7, 5.1) | 51 | 4.4 (3.7, 5.0) | 47 | 4.8 (4.1, 5.5) |
| Postintervention | 43 | 3.8 (3.1, 4.5) | 47 | 4.4 (3.7, 5.0) | 48 | 3.5 (2.9, 4.2) | 43 | 4.0 (3.3, 4.7) |
| Change | 49 | -0.8 (-1.6, 0.0) | 49 | 0.0 (-0.9, 0.8) | 52 | -0.8 (-1.6, -0.1) | 47 | -0.8 (-1.6, 0.1) |
| *Hepatokines* | | | | | | | | |
| Angiopoietin–like 3 (ng/mL) | | | | | | | | |
| Preintervention | 47 | 455.1 (416.9, 493.3) | 47 | 445.6 (407.4, 483.9) | 50 | 419.7 (382.6, 456.8) | 46 | 393.3 (354.6, 432.0) |
| Postintervention | 43 | 351.0 (311.2, 390.8) | 47 | 369.3 (331.0, 407.5) | 48 | 358.5 (320.8, 396.3) | 43 | 331.8 (291.9, 371.6) |
| Change | 49 | -104.1 (-150.6, -57.6) * | 49 | -76.4 (-121.5, -31.2) * | 52 | -61.2 (-105.6, -16.8) * | 47 | -61.5 (-108.1, -15.0) * |
| β–Klotho (ng/mL) | | | | | | | | |

| | | | | | | | | |
|---|---|---|---|---|---|---|---|---|
| Preintervention | 47 | 12.6 (11.1, 14.1) | 45 | 10.9 (9.4, 12.4) | 51 | 11.0 (9.6, 12.5) | 47 | 11.5 (10.0, 13.0) |
| Postintervention | 43 | 10.6 (9.1, 12.1) | 47 | 9.6 (8.1, 11.1) | 47 | 9.8 (8.4, 11.3) | 43 | 11.1 (9.5, 12.6) |
| Change | 49 | -2.1 (-3.2, -0.9) * | 49 | -1.3 (-2.5, -0.1) * | 52 | -1.2 (-2.3, -0.1) * | 47 | -0.5 (-1.6, 0.7) |
| Clusterin (μg/mL) | | | | | | | | |
| Preintervention | 47 | 228.7 (213.1, 244.3) | 47 | 237.9 (222.3, 253.6) | 50 | 231.8 (216.6, 246.9) | 46 | 228.8 (213.0, 244.6) |
| Postintervention | 43 | 208.5 (192.2, 224.8) | 47 | 216.3 (200.7, 232.0) | 48 | 210.1 (194.6, 225.5) | 43 | 204.3 (188.0, 220.6) |
| Change | 49 | -20.2 (-42.8, 2.5) * | 49 | -21.6 (-43.8, 0.5) * | 52 | -21.7 (-43.4, -0.0) * | 47 | -24.5 (-47.3, -1.7) * |
| Follistatin (ng/mL) | | | | | | | | |
| Preintervention | 47 | 1.02 (0.91, 1.14) | 46 | 1.03 (0.92, 1.14) | 49 | 1.01 (0.90, 1.12) | 47 | 0.94 (0.82, 1.05) |
| Postintervention | 43 | 0.92 (0.81, 1.04) | 47 | 0.91 (0.80, 1.03) | 48 | 0.92 (0.80, 1.03) | 43 | 0.85 (0.73, 0.96) |
| Change | 49 | -0.10 (-0.19, -0.01) | 48 | -0.12 (-0.20, -0.03) * | 50 | -0.10 (-0.18, -0.01) * | 47 | -0.09 (-0.17, -0.00) * |
| Sex Hormone-Binding Globulin (nmol/L) | | | | | | | | |
| Preintervention | 44 | 25.9 (20.5, 31.3) | 47 | 22.3 (17.1, 27.5) | 49 | 24.1 (18.9, 29.2) | 45 | 20.5 (15.1, 25.8) |
| Postintervention | 42 | 21.2 (15.7, 26.7) | 43 | 24.6 (19.1, 30.1) | 47 | 19.0 (13.8, 24.2) | 42 | 22.5 (17.0, 28.0) |
| Change | 48 | -4.8 (-12.4, 2.9) | 48 | 2.3 (-5.2, 9.8) | 51 | -5.0 (-12.3, 2.2) | 47 | 2.0 (-5.6, 9.6) |
| *Adipo-hepatokines* | | | | | | | | |
| Angiopoietin–like 4 (ng/mL) | | | | | | | | |
| Preintervention | 47 | 1.28 (0.89, 1.67) | 46 | 0.56 (0.16, 0.95) | 49 | 0.81 (0.43, 1.20) | 47 | 0.84 (0.45, 1.24) |
| Postintervention | 43 | 0.96 (0.56, 1.35) | 47 | 0.46 (0.07, 0.86) | 48 | 0.76 (0.38, 1.15) | 42 | 0.68 (0.28, 1.08) |
| Change | 49 | -0.32 (-0.56, -0.08) | 48 | -0.09 (-0.32, 0.14) * | 50 | -0.05 (-0.28, 0.17) | 47 | -0.17 (-0.40, 0.07) |
| Dipeptidyl Peptidase IV (μg/mL) | | | | | | | | |
| Preintervention | 47 | 1.07 (0.98, 1.15) | 47 | 1.04 (0.95, 1.13) | 51 | 1.01 (0.93, 1.10) | 46 | 0.96 (0.88, 1.05) |
| Postintervention | 43 | 0.85 (0.76, 0.94) | 47 | 0.79 (0.70, 0.87) | 48 | 0.86 (0.77, 0.94) | 43 | 0.80 (0.71, 0.89) |
| Change | 49 | -0.22 (-0.31, -0.12) * | 49 | -0.25 (-0.34, -0.16) * | 52 | -0.16 (-0.25, -0.07) * | 47 | -0.16 (-0.26, -0.07) * |
| Growth Differentiation Factor 15 (ng/mL) | | | | | | | | |
| Preintervention | 47 | 0.28 (0.26, 0.31) | 47 | 0.29 (0.27, 0.32) | 50 | 0.29 (0.27, 0.32) | 47 | 0.25 (0.22, 0.28) |
| Postintervention | 43 | 0.27 (0.24, 0.30) | 47 | 0.27 (0.24, 0.29) | 48 | 0.28 (0.25, 0.30) | 43 | 0.25 (0.22, 0.27) |
| Change | 49 | -0.01 (-0.03, 0.00) * | 49 | -0.03 (-0.04, -0.01) * | 51 | -0.02 (-0.03, -0.00) * | 47 | 0.00 (-0.02, 0.02) |
| Retinol-Binding Protein 4 (μg/mL) | | | | | | | | |
| Preintervention | 47 | 35.6 (31.8, 39.5) | 47 | 36.9 (33.0, 40.8) | 50 | 35.8 (32.1, 39.6) | 46 | 34.8 (30.9, 38.7) |
| Postintervention | 43 | 25.8 (21.8, 29.9) | 47 | 26.1 (22.2, 30.0) | 48 | 24.8 (21.0, 28.7) | 43 | 25.0 (21.0, 29.1) |
| Change | 49 | -9.8 (-14.6, -5.0) * | 49 | -10.8 (-15.5, -6.2) * | 52 | -11.0 (-15.6, -6.4) * | 47 | -9.8 (-14.6, -5.0) * |
| *Dietary intake* | | | | | | | | |
| Energy intake (kcal/day) | | | | | | | | |
| Preintervention | 49 | 2034 (1910, 2158) | 49 | 2024 (1899, 2148) | 52 | 1874 (1754, 1995) | 47 | 1995 (1868, 2122) |
| Postintervention | 46 | 1807 (1679, 1935) | 48 | 1492 (1367, 1618) | 48 | 1503 (1378, 1628) | 43 | 1525 (1393, 1657) |
| Change | 49 | -227 (-368, -86) * | 49 | -531 (-671, -392) * | 52 | -372 (-510, -234) * | 47 | -470 (-616, -324) * |
| Carbohydrate intake (% EI) | | | | | | | | |
| Preintervention | 49 | 36 (33, 38) | 49 | 35 (33, 37) | 52 | 35 (33, 37) | 47 | 37 (35, 40) |
| Postintervention | 46 | 35 (33, 38) | 48 | 35 (33, 38) | 48 | 35 (33, 37) | 43 | 37 (34, 39) |
| Change | 49 | 0 (-3, 2) | 49 | 1 (-2, 3) | 52 | 0 (-2, 2) | 47 | -1 (-3, 2) |
| Fat intake (% EI) | | | | | | | | |
| Preintervention | 49 | 43 (41, 45) | 49 | 44 (42, 46) | 52 | 45 (43, 47) | 47 | 43 (41, 45) |
| Postintervention | 46 | 43 (41, 45) | 48 | 43 (41, 45) | 48 | 43 (41, 45) | 43 | 42 (40, 44) |
| Change | 49 | 0 (-3, 2) | 49 | -1 (-4, 1) | 52 | -2 (-4, 1) | 47 | -1 (-4, 1) |
| Protein intake (% EI) | | | | | | | | |
| Preintervention | 49 | 18 (17, 19) | 49 | 17 (17, 18) | 52 | 18 (17, 19) | 47 | 17 (16, 18) |
| Postintervention | 46 | 19 (18, 20) | 48 | 18 (17, 19) | 48 | 19 (18, 20) | 43 | 19 (18, 20) |
| Change | 49 | 1 (0, 2) | 49 | 1 (0, 2) | 52 | 1 (0, 2) * | 47 | 2 (0, 3) * |
| Alcohol intake (% EI) | | | | | | | | |
| Preintervention | 49 | 3 (2, 4) | 49 | 3 (2, 4) | 52 | 2 (1, 3) | 47 | 2 (1, 3) |
| Postintervention | 46 | 3 (2, 4) | 48 | 3 (2, 4) | 48 | 3 (2, 4) | 43 | 3 (2, 4) |
| Change | 49 | 0 (-1, 1) | 49 | 0 (-1, 1) | 52 | 1 (0, 2) | 47 | 0 (-1, 1) |

Data are presented as estimated marginal means with 95% confidence intervals (CIs) from linear mixed models. These are model-derived estimates, not raw means, which explains any differences with baseline values in Table 1. Asterisks (*) represent statistically significant within-group changes as determined by linear mixed model contrasts ($P < 0.05$). No adjustments were made for multiple comparisons within groups. Hepatic fat fraction was assessed by magnetic resonance imaging. Liver stiffness, attenuation imaging coefficient (ATI), and viscosity were assessed using elastography. Adipokines, hepatokines, and adipo-hepatokines were log10-transformed for statistical analyses; however, results are presented in original units to improve interpretability. As a result, some CIs may appear to include zero or not in the original scale, although all analyses were performed on the log10-transformed data. Dietary intake was estimated from three non-consecutive 24–hour dietary recalls, two on working days and one on a non-working day. Changes were calculated as postintervention minus preintervention values. The number of participants with valid data at each time point (Preintervention, Postintervention) and in the final model (Change) is reported by group for each variable. "Change" reflects the total sample included in the linear mixed models, which account for missing data and may include participants with only Preintervention, only Postintervention, or both measurements. ALT, alanine aminotransferase; ALP, alkaline phosphatase; GGT, γ–glutamyltransferase; TRE, time–restricted eating; UC, usual care.

**Table S2**. Associations between changes in hepatic fat fraction and changes in adipokines, hepatokines, adipo-hepatokines, and insulin resistance after the 12-week intervention in all sample (i.e., including usual care, early time-restricted eating [TRE], late TRE, and self-selected TRE groups), and in TRE groups alone.

| | **All sample** | | **TRE groups alone** | |
|---|---|---|---|---|
| | $r_{rm}$ (95% CI) | *P* value | $r_{rm}$ (95% CI) | *P* value |
| *Adipokines* | | | | |
| Adiponectin (µg/mL) | 0.033 (-0.127, 0.191) | 0.684 | -0.006 (-0.189, 0.177) | 0.947 |
| Interleukin–6 (pg/mL) | -0.028 (-0.187, 0.133) | 0.733 | -0.029 (-0.213, 0.158) | 0.763 |
| Omentin (ng/mL) | 0.104 (-0.056, 0.260) | 0.202 | 0.078 (-0.107, 0.259) | 0.407 |
| Resistin (ng/mL) | 0.024 (-0.136, 0.182) | 0.773 | 0.016 (-0.167, 0.197) | 0.868 |
| *Hepatokines* | | | | |
| Angiopoietin–like 3 (ng/mL) | 0.263 (0.107, 0.405) | **0.001** | 0.242 (0.061, 0.408) | **0.009** |
| β–Klotho (ng/mL) | 0.274 (0.119, 0.416) | **0.001** | 0.288 (0.109, 0.449) | **0.002** |
| Clusterin (µg/mL) | 0.264 (0.109, 0.407) | **0.001** | 0.222 (0.039, 0.390) | **0.018** |
| Follistatin (ng/mL) | 0.264 (0.109, 0.405) | **0.001** | 0.302 (0.127, 0.459) | **0.001** |
| Sex Hormone-Binding Globulin (nmol/L) | 0.087 (-0.078, 0.248) | 0.300 | 0.040 (-0.151, 0.227) | 0.684 |
| *Adipo-hepatokines* | | | | |
| Angiopoietin–like 4 (ng/mL) | -0.072 (-0.228, 0.087) | 0.375 | -0.065 (-0.244, 0.119) | 0.488 |
| Dipeptidyl Peptidase IV (µg/mL) | 0.307 (0.156, 0.445) | **< 0.001** | 0.263 (0.084, 0.426) | **0.005** |
| Growth Differentiation Factor 15 (ng/mL) | 0.033 (-0.126, 0.191) | 0.685 | 0.030 (-0.153, 0.211) | 0.747 |
| Retinol-Binding Protein 4 (µg/mL) | 0.402 (0.258, 0.527) | **< 0.001** | 0.387 (0.219, 0.533) | **< 0.001** |
| *Insulin resistance* | | | | |
| HOMA-IR | 0.416 (0.276, 0.540) | **< 0.001** | 0.360 (0.188, 0.511) | **< 0.001** |

Repeated measures correlation ($r_{rm}$) was used to assess longitudinal associations between within-individual changes in hepatic fat fraction and changes in adipokines, hepatokines, adipohepatokines, and insulin resistance. Positive $r_{rm}$ values indicate that reductions in hepatic fat fraction were associated with decreases in the respective biomarker or insulin resistance, whereas negative values suggest that reductions in hepatic fat fraction are associated with increases in the respective biomarker or insulin resistance. Adipokines, hepatokines, and adipo-hepatokines were log10-transformed for the present statistical analyses. Confidence intervals (CIs) were calculated using an analytic approach with a 95% confidence level. Significant associations are indicated in bold ($P < 0.05$). HOMA-IR, homeostasis model assessment of insulin resistance.

**Table S3**. Alpha diversity indices of gut microbiota before and after the intervention in the usual care (UC), early time–restricted eating (TRE), late TRE, and self–selected TRE groups.

| Endpoint | UC (n = 35) | | | Early TRE (n = 36) | | | Late TRE (n = 36) | | | Self-selected TRE (n = 31) | | |
|---|---|---|---|---|---|---|---|---|---|---|---|---|
| | Before | After | *P* | Before | After | *P* | Before | After | *P* | Before | After | *P* |
| **Shannon entropy** | 5.88 (5.52 – 6.16) | 5.66 (5.28 – 6.10) | 0.20 | 5.97 (5.55 – 6.16) | 5.78 (5.44 – 6.08) | 0.43 | 5.79 (5.24 – 6.18) | 5.95 (5.51 – 6.25) | 0.90 | 6.00 (5.61 – 6.41) | 6.00 (5.57 – 6.20) | 0.72 |
| **Simpson index** | 0.96 (0.95 – 0.97) | 0.95 (0.93 – 0.97) | **0.02** | 0.97 (0.95 – 0.97) | 0.96 (0.94 – 0.97) | 0.25 | 0.96 (0.94 – 0.97) | 0.96 (0.94 – 0.97) | 0.66 | 0.97 (0.95 – 0.98) | 0.97 (0.95 – 0.98) | 0.99 |
| **Observed features** | 321 (78) | 340 (103) | 0.27 | 315 (92) | 346 (74) | 0.06 | 325 (102) | 355 (91) | 0.10 | 347 (108) | 342 (92) | 0.79 |
| **Pielou evenness** | 0.70 (0.68 – 0.73) | 0.68 (0.65 – 0.70) | **0.02** | 0.70 (0.67 – 0.74) | 0.69 (0.65 – 0.72) | 0.10 | 0.69 (0.66 – 0.73) | 0.69 (0.65 – 0.73) | 0.26 | 0.72 (0.68 – 0.74) | 0.71 (0.68 – 0.74) | 0.87 |

Data are presented as mean (standard deviation) when normally distributed, or medians (first quartile – third quartile) when not. Within-group comparisons were performed using the t paired t-test or the Wilcoxon signed-rank test, depending on whether the data were normally distributed or not, respectively. Significant differences are indicated in bold ($P < 0.05$).

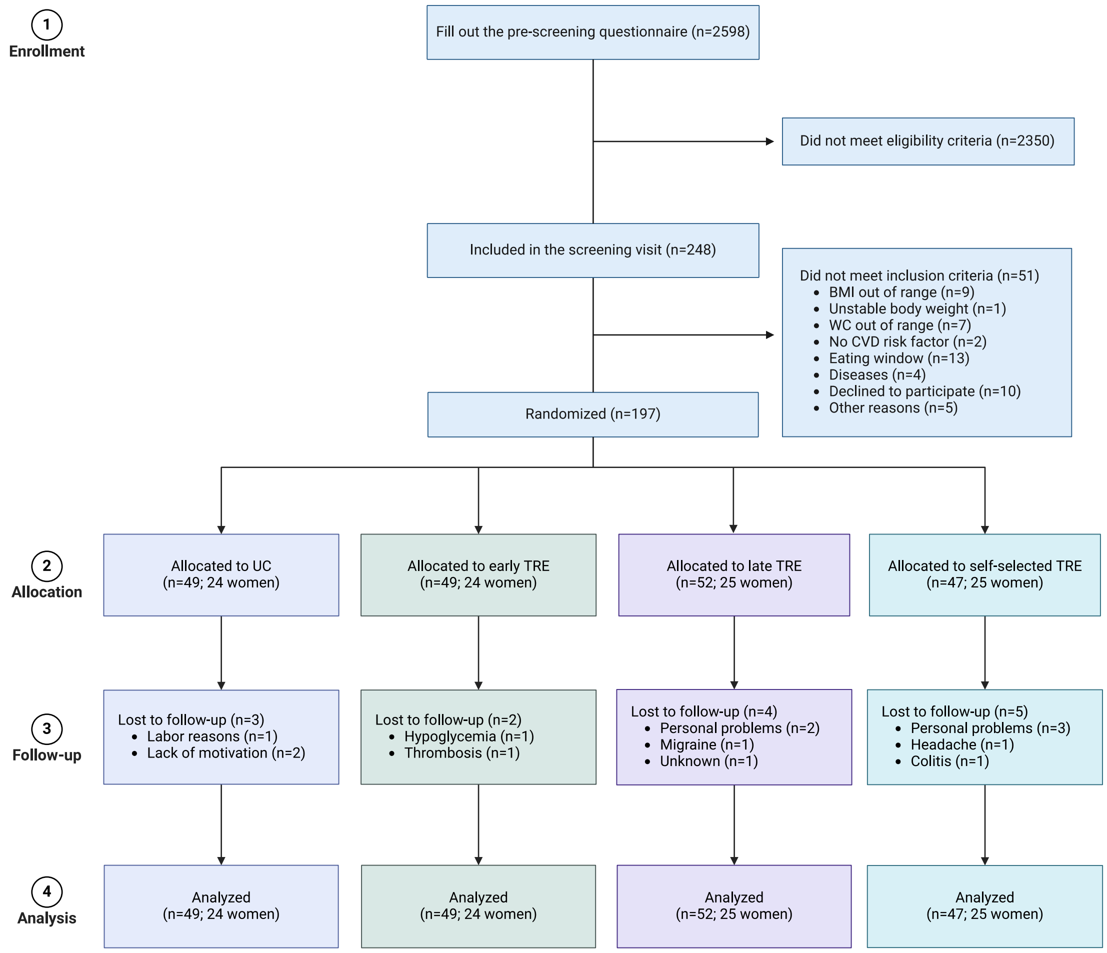

**Figure S1.** Study flow chart. Usual care (UC), early time–restricted eating (TRE), late TRE, or self–selected TRE groups. WC, waist circumference; CVD, cardiovascular; BMI, body mass index. Created in BioRender. Ruiz, J. (2024) BioRender.com/t37h141

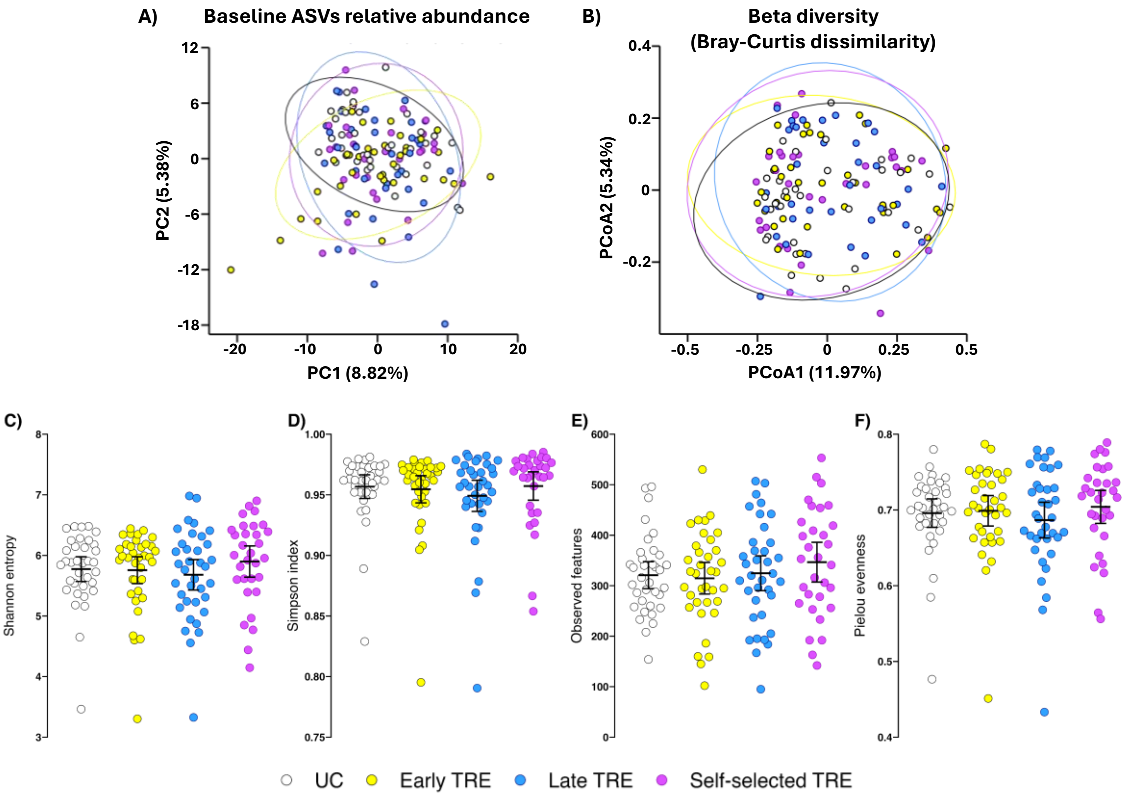

**Figure S2**. Baseline gut microbiota composition and diversity in the the usual care (UC), early time–restricted eating (TRE), late TRE, and self–selected TRE groups. **(A)** Principal component analysis (PCA) plot of the baseline relative abundance of the amplicon sequence variants (ASVs) detected through 16S rRNA sequencing ($P$ = 0.29). **(B)** Principal coordinate analysis (PCoA) plot based on Bray-Curtis distance of baseline gut microbiota composition ($P$ = 0.30). **(C-F)** Comparison of baseline alpha-diversity metrics between groups: **(C)** Shannon entropy ($P$ = 0.42), **(D)** Simpson index ($P$ = 0.45), **(E)** observed features ($P$ = 0.63), and **(F)** Pielou evenness ($P$ = 0.65). Statistical analyses were performed using MANOVA and PERMANOVA for PCA and PCoA analyses, respectively, and the Kruskal-Wallis test for the comparisons between groups for alpha-diversity indices. Significant differences were set at $P < 0.05$.

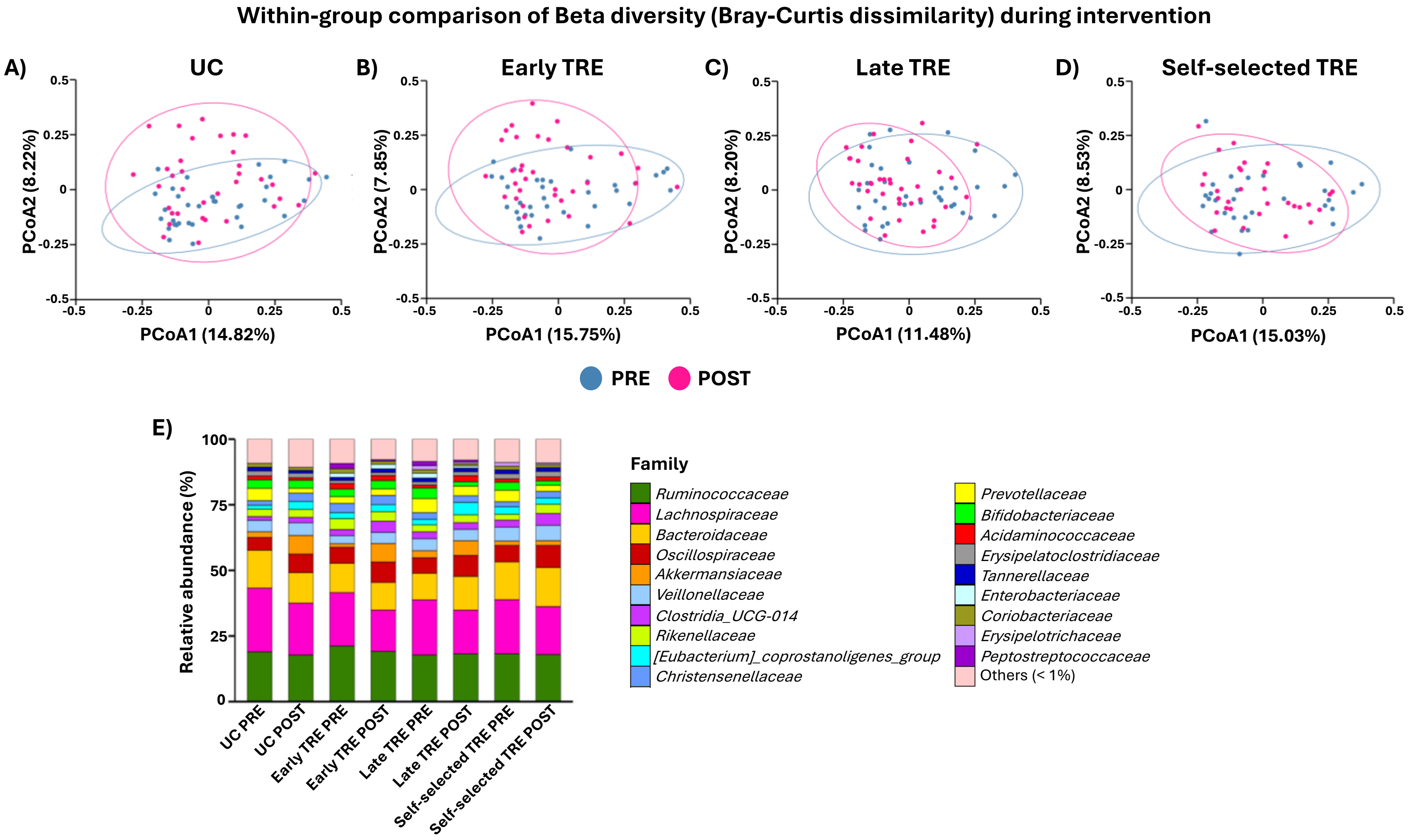

**Figure S3**. **(A-D)** PCoA plots representing within-group comparisons of beta-diversity (Bray-Curtis dissimilarity) at the two-time points of the intervention, PRE (blue) and POST (pink), for the usual care (UC; A; $P = 0.26$), early time–restricted eating (TRE; B; $P = 0.10$), late TRE (C; $P = 0.19$), and self–selected TRE (D; $P = 0.76$) groups. Statistical analyses for PCoA were performed using PERMANOVA. **(E)** Microbial taxonomic composition at the family level in fecal samples for the PRE and POST time points for each intervention group. The bars show the mean proportion of relative abundance. The "Others" category in the graph represents the aggrupation of bacterial groups with less than 1% relative abundance. Changes in bacterial composition were evaluated through MaAsLin3 algorithm considering the assigned group, assessment time, and their interactions terms with sex as a covariate. No statistically significant differences were detected after applying Benjamini-Hochberg False Discovery Rate (FDR). Significant differences were set at $P < 0.05$.

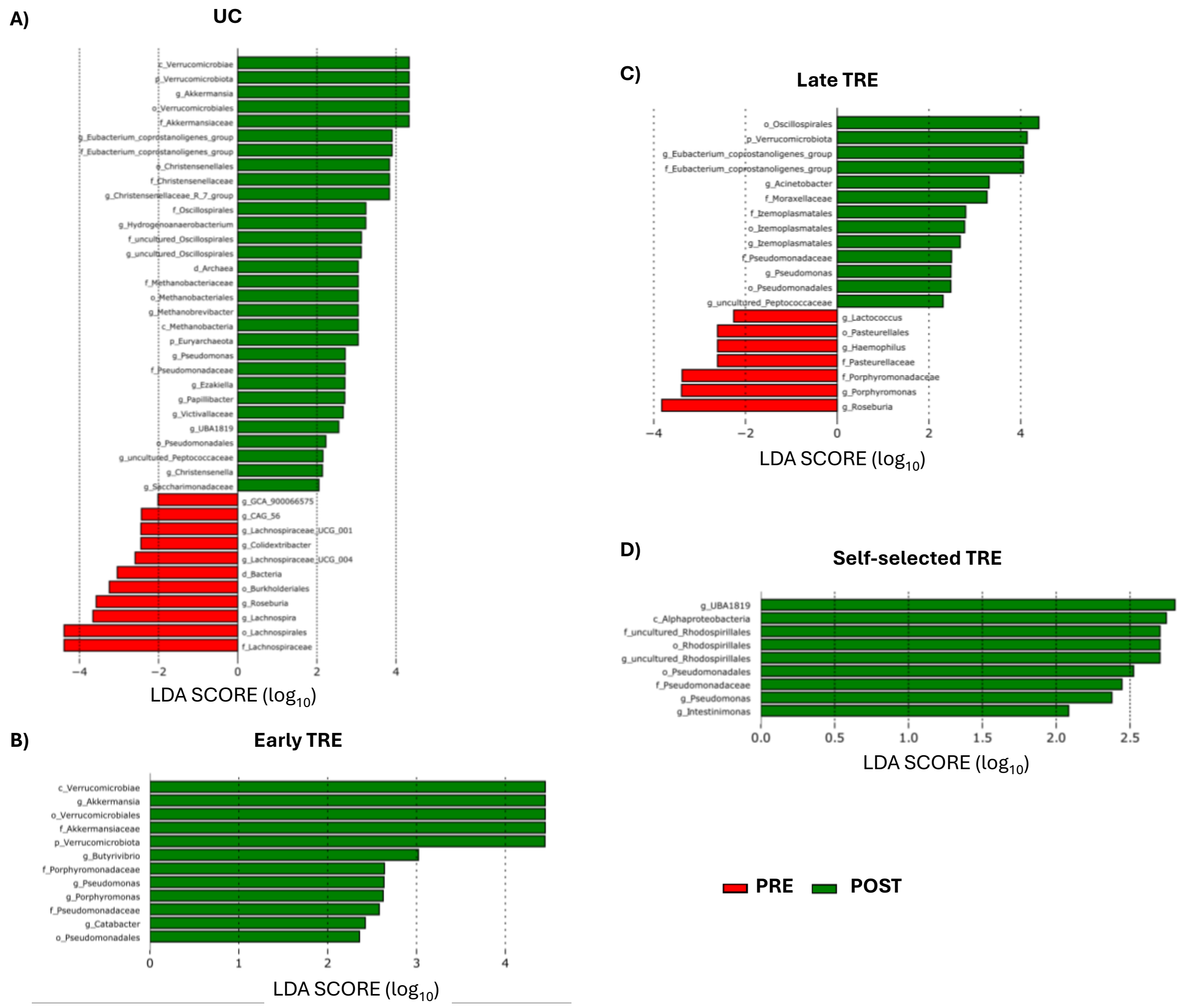

**Figure S4**. Linear discriminant analysis effect size (LEfSe) for taxa biomarker discovery between preintervention (PRE) and postintervention (POST) within the usual care (UC; A), early time–restricted eating (TRE; B), late TRE (C), and self–selected TRE (D) groups. Histograms of the Linear Discriminant Analysis (LDA) scores show features that are differentially abundant between PRE (red) and POST (green) in each intervention group. The LDA score threshold was set to 2.0, and significant differences were set at $P < 0.05$.

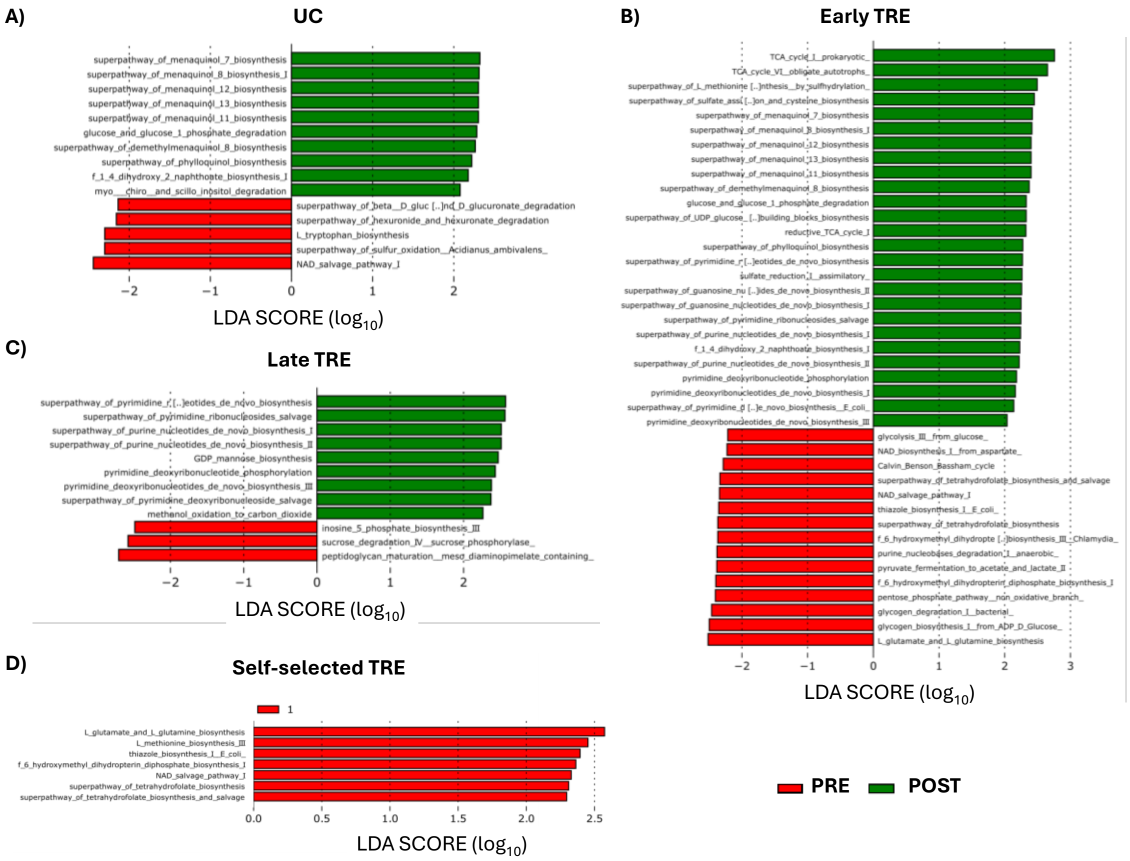

**Figure S5**. Linear discriminant analysis effect size (LEfSe) for metabolic pathway biomarker discovery between PRE and POST time points within the usual care (UC; A), early time–restricted eating (TRE; B), late TRE (C), and self–selected TRE (D) groups. Histograms of the Linear Discriminant Analysis (LDA) scores show differentially abundant bacterial pathways between preintervention (PRE; red) and postintervention (POST; green) in each intervention group. The LDA score threshold was set to 2.0, and significant differences were set at $P < 0.05$.

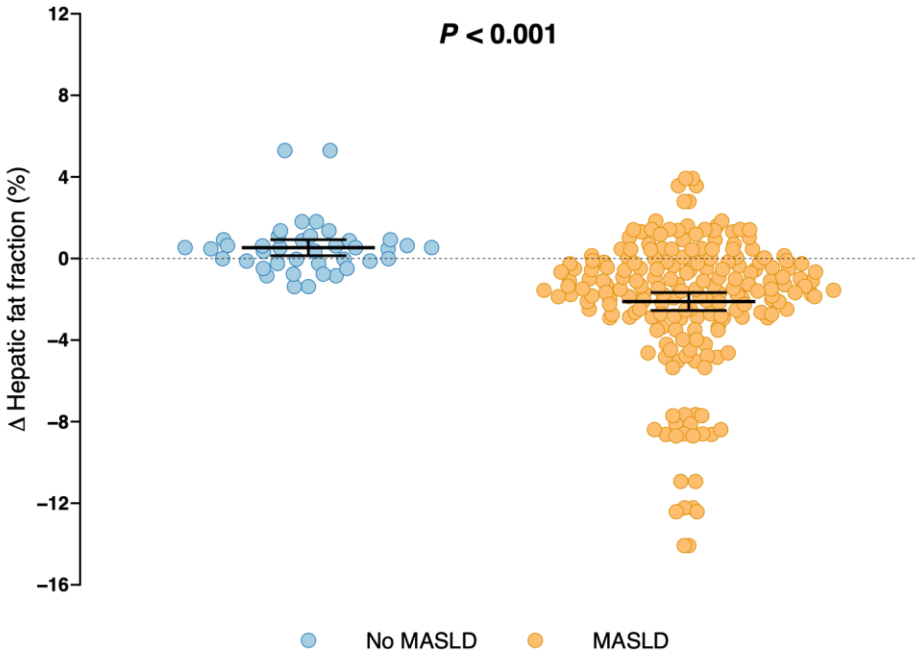

**Figure S6.** Changes in hepatic fat fraction in participants from the time-restricted eating groups, stratified by the presence of metabolic dysfunction-associated steatotic liver disease (MASLD) at baseline (defined as ≥5% hepatic fat fraction) compared to those without the condition (defined as <5% hepatic fat fraction). Data are raw means with 95% confidence interval. Between-group differences (No MASLD vs. MASLD) were evaluated using linear mixed-effects models with repeated measures adjusted for sex. Statistically significant differences are displayed in bold ($P < 0.05$). Hepatic fat fraction was assessed by magnetic resonance imaging. Changes were calculated as postintervention minus preintervention values.

**Table S4.** Baseline characteristics of participants classified as non-responders (i.e., body weight loss <5%) and responders (i.e., body weight loss ≥5%).

| | **Non-responders (n = 117)** | **Responders (n = 69)** |
|---|---|---|
| Age (years) | 46.9 (6.6) | 46.9 (5.8) |
| Women, No. (%) | 57 (48.7) | 31 (44.9) |
| Weight (kg) | 94.3 (16.2) | 97.6 (11.6) |
| Height (cm) | 169.4 (9.4) | 170.8 (8.4) |
| Body mass index (kg/m$^2$) | 32.7 (3.9) | 33.4 (2.7) |
| Waist circumference (cm) | 100.9 (10.4) | 103.7 (9.1) |
| Hip circumference (cm) | 113.9 (8.6) | 115.1 (7.3) |
| Waist to hip ratio | 0.9 (0.1) | 0.9 (0.1) |
| *Liver parameters* | | |
| Hepatic fat fraction (%) | 7.5 (5.6 – 11.5) | 8.6 (5.7 – 11.6) |
| MASLD prevalence, No. (%)[a] | 93 (82.3) | 53 (86.9) |
| Stage of hepatic steatosis[a] | | |
| S0, No. (%) | 20 (17.7) | 8 (13.1) |
| S1, No. (%) | 79 (69.9) | 42 (68.9) |
| S2, No. (%) | 12 (10.6) | 8 (13.1) |
| S3, No. (%) | 2 (1.8) | 3 (4.9) |
| Liver stiffness (kPa) | 6.7 (5.8 – 7.6) | 6.3 (5.2 – 7.5) |
| ATI (dB/cm/MHz) | 0.70 (0.64 – 0.76) | 0.70 (0.64 – 0.76) |
| Viscosity (m/s/kHz) | 10.4 (9.2 – 12.3) | 11.2 (9.3 – 12.6) |
| ALT (U/L) | 23 (17 – 32) | 29 (21 – 39) |
| GGT (U/L) | 25 (18 – 36) | 26 (19 – 42) |
| ALP (U/L) | 61 (53 – 75) | 66 (57 – 78) |
| *Adipokines* | | |
| Adiponectin (µg/mL) | 6.3 (4.2 – 9.3) | 5.6 (3.4 – 7.1) |
| Interleukin–6 (pg/mL) | 2.1 (1.4 – 2.9) | 1.9 (1.5 – 2.6) |
| Omentin (ng/mL) | 49.4 (35.7 – 65.3) | 53.2 (35.0 – 69.7) |
| Resistin (ng/mL) | 4.3 (2.8 – 6.0) | 3.8 (2.6 – 4.8) |
| *Hepatokines* | | |
| Angiopoietin–like 3 (ng/mL) | 386.2 (312.9 – 501.3) | 412.0 (319.5 – 489.8) |
| β–Klotho (ng/mL) | 10.9 (9.7 – 11.9) | 10.6 (9.9 – 11.9) |
| Clusterin (µg/mL) | 222.0 (195.5 – 260.7) | 231.6 (203.3 – 261.3) |
| Follistatin (ng/mL) | 0.98 (0.78 – 1.17) | 0.90 (0.73 – 1.13) |
| Sex Hormone–Binding Globulin (nmol/L) | 19.6 (14.9 – 27.3) | 17.5 (10.6 – 28.5) |
| *Adipo–hepatokines* | | |
| Angiopoietin–like 4 (ng/mL) | 0.28 (0.17 – 1.18) | 0.21 (0.15 – 0.70) |
| Dipeptidyl Peptidase IV (µg/mL) | 0.96 (0.79 – 1.24) | 0.96 (0.85 – 1.13) |
| Growth Differentiation Factor 15 (ng/mL) | 0.27 (0.23 – 0.34) | 0.25 (0.22 – 0.30) |
| Retinol–Binding Protein 4 (µg/mL) | 34.1 (24.3 – 46.9) | 29.7 (21.9 – 43.4) |
| *Dietary intake* | | |
| Energy intake (kcal/day) | 1941 (1620 – 2242) | 1960 (1718 – 2271) |
| Carbohydrate intake (% EI) | 36 (31 – 40) | 36 (32 – 39) |
| Fat intake (% EI) | 17 (15 – 20) | 17 (15 – 20) |
| Protein intake (% EI) | 44 (40 – 47) | 44 (40 – 48) |
| Alcohol intake (% EI) | 2 (0 – 4) | 1 (0 – 3) |
| *Cardiovascular risk factors* | | |
| Systolic BP (mm Hg) | 123 (14) | 124 (14) |
| Diastolic BP (mm Hg) | 81 (10) | 80 (11) |
| Fasting glucose (mg/dL) | 92 (88 – 98) | 94 (90 – 100) |
| Fasting insulin (mU/L) | 10.2 (7.6 – 13.5) | 10.9 (8.8 – 15.1) |
| HOMA–IR | 2.46 (1.64 – 3.18) | 2.53 (1.99 – 3.80) |
| HbA1c (%) | 5.3 (5.1 – 5.5) | 5.4 (5.2 – 5.6) |
| Total cholesterol (mg/dL) | 210 (184 – 228) | 207 (186 – 231) |
| HDL–C (mg/dL) | 54 (45 – 61) | 51 (45 – 57) |
| LDL–C (mg/dL) | 127 (111 – 147) | 136 (116 – 156) |

| | | |
|---|---|---|
| Triglycerides (mg/dL) | 123 (82 – 159) | 114 (90 – 149) |

Data are presented as mean (standard deviation) when normally distributed, median (first quartile – third quartile) when not normally distributed, or as frequency (associated percentage). Hepatic fat fraction was assessed by magnetic resonance imaging (MRI). Metabolic dysfunction-associated steatotic liver disease (MASLD) prevalence was defined as the number and percentage of participants with hepatic fat fraction ≥5%. Stages of hepatic steatosis were defined as follows: S0: no steatosis [hepatic fat fraction <5%]; S1: mild steatosis [hepatic fat fraction ≥5% and <15%]; S2: moderate steatosis [hepatic fat fraction ≥15% and <25%]; S3: severe steatosis [hepatic fat fraction ≥25%].[23] Liver stiffness, attenuation imaging coefficient (ATI), and viscosity were assessed using elastography. Dietary intake was estimated from three non-consecutive 24–hour dietary recalls, two on working days and one on a non-working day. No statistically significant differences were observed at baseline for any outcome across groups (analysis of variance with Benjamini–Hochberg adjustment; all $P \geq 0.31$). ALT, alanine aminotransferase; ALP, alkaline phosphatase; APOA1, apolipoprotein A; APOB, apolipoprotein B; BP, blood pressure; EI, energy intake; GGT, γ–glutamyltransferase; HbA1c, glycated hemoglobin; HDL–C, high–density lipoprotein cholesterol; HOMA–IR, homeostasis model assessment of insulin resistance; LDL–C, low–density lipoprotein cholesterol; TRE, time–restricted eating; UC, usual care. [a]Calculated based on the maximum number of participants with available hepatic fat fraction data.

**Table S5**. Liver parameters, adipokines, hepatokines, adipo-hepatokines, and dietary intake endpoints at baseline and after the 12–week intervention in non-responders (i.e., body weight loss <5%) and responders (i.e., body weight loss ≥5%).

| **Endpoint** | **Non-responders (n = 117)** | | **Responders (n = 69)** | | **Responders vs. non-responders** | |
|---|---|---|---|---|---|---|
| | n | Mean (95% CI) | n | Mean (95% CI) | Mean difference (95% CI) | *P* value |
| *Liver parameters* | | | | | | |
| Hepatic fat fraction (%) | | | | | | |
| Preintervention | 113 | 9.4 (8.5, 10.3) | 61 | 10.2 (9.0, 11.5) | | |
| Postintervention | 103 | 8.9 (8.0, 9.9) | 65 | 7.2 (5.9, 8.4) | | |
| Change | 115 | -0.5 (-1.1, 0.1) | 67 | -3.1 (-3.8, -2.3) * | -2.6 (-3.6, -1.6) | **<0.001** |
| Liver stiffness (kPa) | | | | | | |
| Preintervention | 84 | 7.1 (6.5, 7.6) | 47 | 7.2 (6.5, 7.9) | | |
| Postintervention | 98 | 6.9 (6.4, 7.4) | 49 | 7.2 (6.5, 7.9) | | |
| Change | 105 | -0.2 (-0.7, 0.3) | 61 | 0.0 (-0.7, 0.8) | 0.2 (-0.7, 1.1) | 0.798 |
| ATI (dB/cm/MHz) | | | | | | |
| Preintervention | 84 | 0.72 (0.70, 0.74) | 50 | 0.72 (0.69, 0.75) | | |
| Postintervention | 98 | 0.68 (0.66, 0.70) | 49 | 0.67 (0.64, 0.70) | | |
| Change | 105 | -0.04 (-0.06, -0.02) * | 63 | -0.05 (-0.08, -0.02) * | -0.01 (-0.05, 0.02) | 0.724 |
| Viscosity (m/s/kHz) | | | | | | |
| Preintervention | 64 | 11.1 (10.4, 11.9) | 27 | 11.3 (10.2, 12.4) | | |
| Postintervention | 66 | 11.2 (10.5, 11.9) | 27 | 11.1 (10.0, 12.2) | | |
| Change | 66 | 0.1 (-0.8, 1.0) | 27 | -0.2 (-1.6, 1.2) | -0.3 (-2.0, 1.3) | 0.831 |
| ALT (U/L) | | | | | | |
| Preintervention | 117 | 27 (25, 30) | 69 | 30 (26, 34) | | |
| Postintervention | 115 | 27 (24, 30) | 67 | 21 (17, 25) | | |
| Change | 117 | 0 (-3, 2) | 69 | -9 (-12, -6) * | -9 (-13, -5) | **<0.001** |
| GGT (U/L) | | | | | | |
| Preintervention | 117 | 38 (29, 47) | 69 | 31 (19, 42) | | |
| Postintervention | 115 | 35 (27, 44) | 67 | 24 (12, 35) | | |
| Change | 117 | -3 ( -7, 1) | 69 | -7 (-12, -2) * | -4 (-11, 2) | 0.400 |
| ALP (U/L) | | | | | | |
| Preintervention | 117 | 65 (62, 68) | 69 | 68 (64, 72) | | |
| Postintervention | 115 | 67 (64, 70) | 67 | 67 (63, 71) | | |
| Change | 117 | 2 (0, 4) | 69 | -1 (-3, 1) | -3 (-6, 0) | 0.206 |
| *Adipokines* | | | | | | |
| Adiponectin (μg/mL) | | | | | | |
| Preintervention | 111 | 7.5 (6.5, 8.4) | 69 | 6.4 (5.2, 7.6) | | |
| Postintervention | 114 | 7.0 (6.1, 7.9) | 67 | 5.2 (4.0, 6.4) | | |
| Change | 117 | -0.5 (-1.3, 0.4) | 69 | -1.2 (-2.3, -0.2) | -0.8 (-2.1, 0.6) | 0.840 |
| Interleukin–6 (pg/mL) | | | | | | |
| Preintervention | 111 | 2.5 (2.2, 2.8) | 66 | 2.2 (1.8, 2.6) | | |
| Postintervention | 115 | 2.2 (1.9, 2.5) | 67 | 2.1 (1.7, 2.4) | | |
| Change | 116 | -0.3 (-0.6, 0.1) * | 68 | -0.2 (-0.6, 0.3) | 0.1 (-0.5, 0.7) | 0.840 |
| Omentin (ng/mL) | | | | | | |
| Preintervention | 111 | 62.7 (51.9, 73.4) | 68 | 58.2 (44.3, 72.2) | | |
| Postintervention | 114 | 65.4 (54.7, 76.1) | 67 | 51.2 (37.2, 65.2) | | |
| Change | 117 | 2.8 ( -2.2, 7.7) | 69 | -7.0 (-13.4, -0.7) * | -9.8 (-17.8, -1.8) | **0.046** |
| Resistin (ng/mL) | | | | | | |
| Preintervention | 112 | 4.9 (4.5, 5.4) | 69 | 4.0 (3.5, 4.6) | | |
| Postintervention | 114 | 4.0 (3.6, 4.5) | 67 | 3.8 (3.2, 4.4) | | |
| Change | 117 | -0.9 (-1.4, -0.4) * | 69 | -0.2 (-0.9, 0.5) | 0.7 (-0.2, 1.5) | 0.348 |
| *Hepatokines* | | | | | | |
| Angiopoietin–like 3 (ng/mL) | | | | | | |
| Preintervention | 111 | 426.4 (401.2, 451.5) | 68 | 438.1 (405.9, 470.4) | | |
| Postintervention | 114 | 367.7 (342.8, 392.5) | 67 | 330.6 (298.1, 363.0) | | |
| Change | 117 | -58.7 ( -87.7, -29.7) * | 69 | -107.5 (-144.9, -70.2) * | -48.8 (-96.2, -1.5) | **0.046** |

| | | | | | | |
|---|---|---|---|---|---|---|
| β–Klotho (ng/mL) | | | | | | |
| Preintervention | 111 | 11.5 (10.6, 12.5) | 68 | 11.2 (10.0, 12.3) | | |
| Postintervention | 113 | 10.5 (9.6, 11.4) | 67 | 9.6 (8.4, 10.8) | | |
| Change | 117 | -1.0 (-1.8, -0.3) * | 69 | -1.5 (-2.5, -0.6) * | -0.5 (-1.7, 0.7) | 0.400 |
| Clusterin (μg/mL) | | | | | | |
| Preintervention | 111 | 231.9 (222.0, 241.9) | 68 | 234.1 (221.4, 246.8) | | |
| Postintervention | 114 | 221.2 (211.3, 231.0) | 67 | 190.8 (178.0, 203.7) | | |
| Change | 117 | -10.8 (-24.8, 3.2) * | 69 | -43.3 (-61.4, -25.2) * | -32.5 (-55.4, -9.6) | **0.025** |
| Follistatin (ng/mL) | | | | | | |
| Preintervention | 112 | 1.02 (0.95, 1.10) | 68 | 0.96 (0.86, 1.05) | | |
| Postintervention | 114 | 0.94 (0.86, 1.01) | 67 | 0.83 (0.74, 0.93) | | |
| Change | 117 | -0.09 (-0.14, -0.03) * | 68 | -0.12 (-0.19, -0.05) * | -0.04 (-0.12, 0.05) | 0.622 |
| Sex Hormone-Binding Globulin (nmol/L) | | | | | | |
| Preintervention | 107 | 22.8 (19.5, 26.2) | 67 | 22.1 (17.8, 26.4) | | |
| Postintervention | 110 | 23.8 (20.5, 27.1) | 64 | 18.1 (13.7, 22.5) | | |
| Change | 116 | 1.0 ( -3.7, 5.7) | 67 | -4.0 (-10.1, 2.0) | -5.0 (-12.7, 2.7) | 0.746 |
| *Adipo-hepatokines* | | | | | | |
| Angiopoietin–like 4 (ng/mL) | | | | | | |
| Preintervention | 112 | 0.89 (0.64, 1.15) | 68 | 0.91 (0.58, 1.25) | | |
| Postintervention | 113 | 0.78 (0.52, 1.04) | 67 | 0.68 (0.35, 1.02) | | |
| Change | 117 | -0.12 (-0.26, 0.03) * | 68 | -0.23 (-0.42, -0.04) | -0.11 (-0.35, 0.13) | 0.400 |
| Dipeptidyl Peptidase IV (μg/mL) | | | | | | |
| Preintervention | 111 | 1.04 (0.99, 1.10) | 69 | 1.00 (0.93, 1.07) | | |
| Postintervention | 114 | 0.87 (0.81, 0.92) | 67 | 0.76 (0.69, 0.83) | | |
| Change | 117 | -0.18 (-0.24, -0.12) * | 69 | -0.24 (-0.32, -0.17) * | -0.06 (-0.16, 0.03) | 0.622 |
| Growth Differentiation Factor 15 (ng/mL) | | | | | | |
| Preintervention | 112 | 0.29 (0.27, 0.31) | 69 | 0.26 (0.24, 0.28) | | |
| Postintervention | 114 | 0.27 (0.26, 0.29) | 67 | 0.25 (0.23, 0.28) | | |
| Change | 117 | -0.02 (-0.03, -0.01) * | 69 | -0.01 (-0.02, 0.01) | 0.01 (-0.01, 0.03) | 0.461 |
| Retinol-Binding Protein 4 (μg/mL) | | | | | | |
| Preintervention | 111 | 37.4 (34.9, 39.9) | 68 | 33.8 (30.6, 37.0) | | |
| Postintervention | 114 | 27.4 (24.9, 29.8) | 67 | 22.3 (19.1, 25.5) | | |
| Change | 117 | -10.0 (-13.0, -7.0) * | 69 | -11.5 (-15.3, -7.7) * | -1.5 (-6.3, 3.4) | 0.371 |
| *Dietary intake* | | | | | | |
| Energy intake (kcal/day) | | | | | | |
| Preintervention | 117 | 1966 (1884, 2048) | 69 | 2022 (1916, 2128) | | |
| Postintervention | 117 | 1622 (1541, 1704) | 68 | 1516 (1409, 1623) | | |
| Change | 117 | -343 (-434, -253) * | 69 | -506 (-624, -389) * | -163 (-311, -15) | 0.131 |
| Carbohydrate intake (% EI) | | | | | | |
| Preintervention | 117 | 35 (34, 37) | 69 | 36 (34, 37) | | |
| Postintervention | 117 | 36 (34, 37) | 68 | 35 (33, 37) | | |
| Change | 117 | 0 (-1, 2) | 69 | 0 (-2, 1) | -1 (-3, 2) | 0.798 |
| Fat intake (% EI) | | | | | | |
| Preintervention | 117 | 44 (42, 45) | 69 | 45 (43, 46) | | |
| Postintervention | 117 | 43 (41, 44) | 68 | 43 (42, 45) | | |
| Change | 117 | -1 (-3, 1) | 69 | -1 (-3, 1) | 0 (-3, 2) | 0.840 |
| Protein intake (% EI) | | | | | | |
| Preintervention | 117 | 18 (17, 18) | 69 | 18 (17, 18) | | |
| Postintervention | 117 | 18 (18, 19) | 68 | 19 (18, 20) | | |
| Change | 117 | 1 (0, 2) * | 69 | 2 (1, 3) * | 1 (0, 2) | 0.400 |
| Alcohol intake (% EI) | | | | | | |
| Preintervention | 117 | 3 (2, 4) | 69 | 2 (1, 3) | | |
| Postintervention | 117 | 3 (3, 4) | 68 | 2 (1, 3) | | |
| Change | 117 | 0 ( 0, 1) | 69 | 0 (-1, 1) | 0 (-1, 1) | 0.840 |

Data for preintervention, postintervention, and change are presented as estimated marginal means with 95% confidence intervals (CIs) from linear mixed models. These are model-derived estimates, not raw means, which explains any differences with baseline values in Table S4. Asterisks (*) represent statistically significant within-group changes as determined by linear mixed model contrasts ($P < 0.05$). Data for between-group differences (non-responders vs. responders) are presented as estimated mean differences and 95% CIs and were evaluated using linear mixed-effects models with repeated measures, adjusted for sex. *P* values were corrected using the Benjamini-Hochberg False Discovery Rate (FDR) method to control for multiple comparisons across outcomes. Differences are reported as responders minus non-responders. Statistically significant differences are displayed in bold (FDR-adjusted *P* < 0.05). Participants classified as non-responders did not achieve clinically significant body weight loss (<5%), while responders achieved ≥5% body weight loss. All participants from usual care (UC), early time-restricted eating (TRE), late TRE, and self-selected TRE groups with available body weight data at both preintervention and postintervention assessments were included in the present analyses. Sample sizes for each category are as follows: UC = 40 non-responders, and 6 responders; early TRE = 29 non-responders, and 20 responders; late TRE = 25 non-responders, and 23 responders; self-selected TRE = 23 non-responders, and 20 responders. Hepatic fat fraction was assessed by magnetic resonance imaging. Liver stiffness, attenuation imaging coefficient (ATI), and viscosity were assessed using elastography. Adipokines, hepatokines, and adipo-hepatokines were log10-transformed for statistical analyses; however, results are presented in original units to improve interpretability. As a result, some confidence intervals may appear to include zero or not in the original scale, although all analyses were performed on the log10-transformed data. Dietary intake was estimated from three non-consecutive 24–hour dietary recalls, two on working days and one on a non-working day. Changes were calculated as postintervention minus preintervention values. The number of participants with valid data at each time point (Preintervention, Postintervention) and in the final model (Change) is reported by group for each variable. "Change" reflects the total sample included in the linear mixed models, which account for missing data and may include participants with only Preintervention, only Postintervention, or both measurements. ALT, alanine aminotransferase; ALP, alkaline phosphatase; EI, energy intake; GGT, γ–glutamyltransferase.

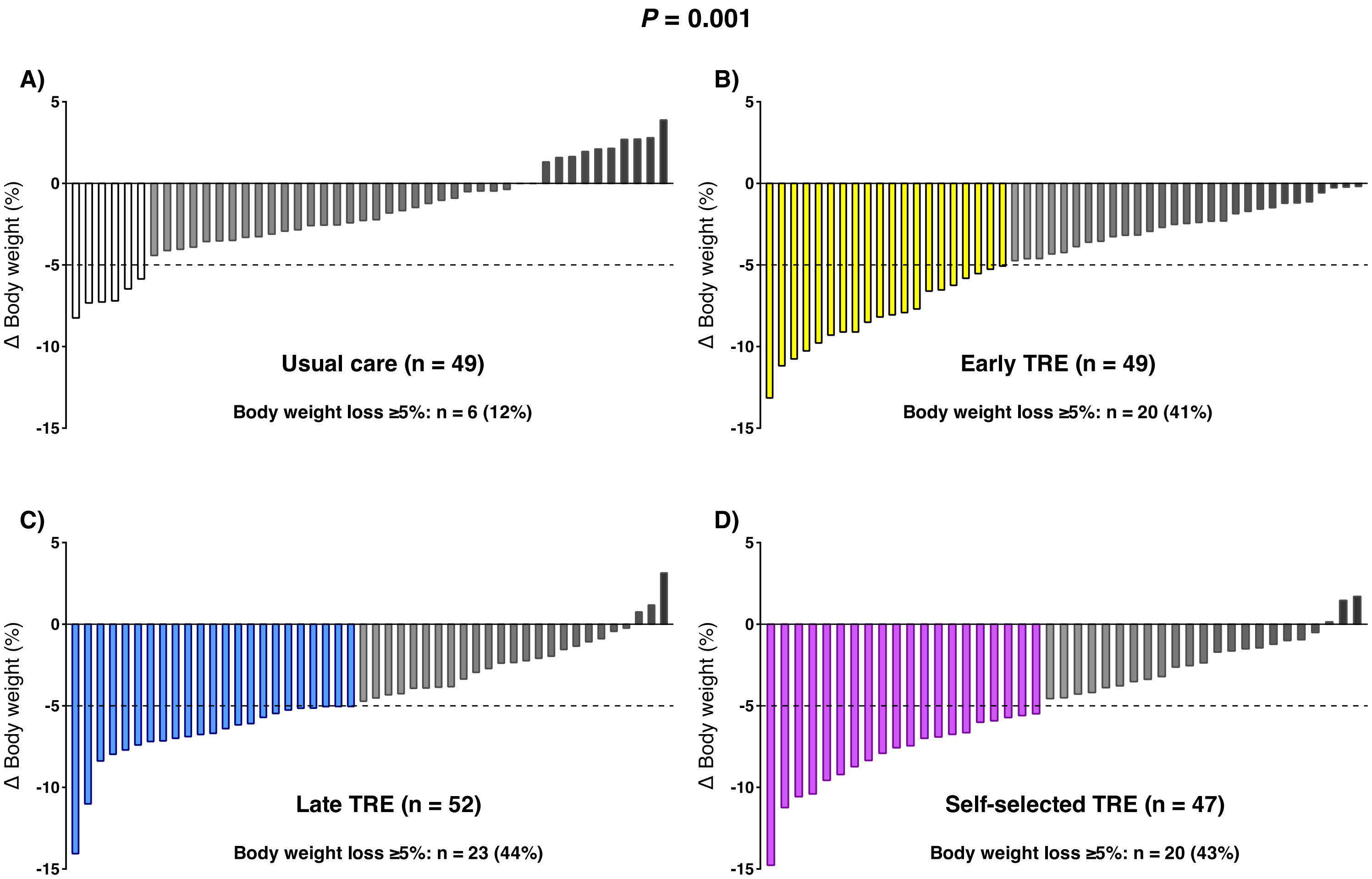

**Figure S7.** Changes in body weight percentage among the usual care (A), early time–restricted eating (TRE; B), late TRE (C), and self–selected TRE (D) groups after the 12-week intervention. The columns represent the raw data of individual study participants. Bars in color indicate participants who achieved clinically meaningful weight loss (≥5%), while gray bars indicate those who did not (<5%). The dotted line represents the 5% weight loss threshold. *P* value from Pearson's chi-square. Body weight percentages indicate changes relative to preintervention values.

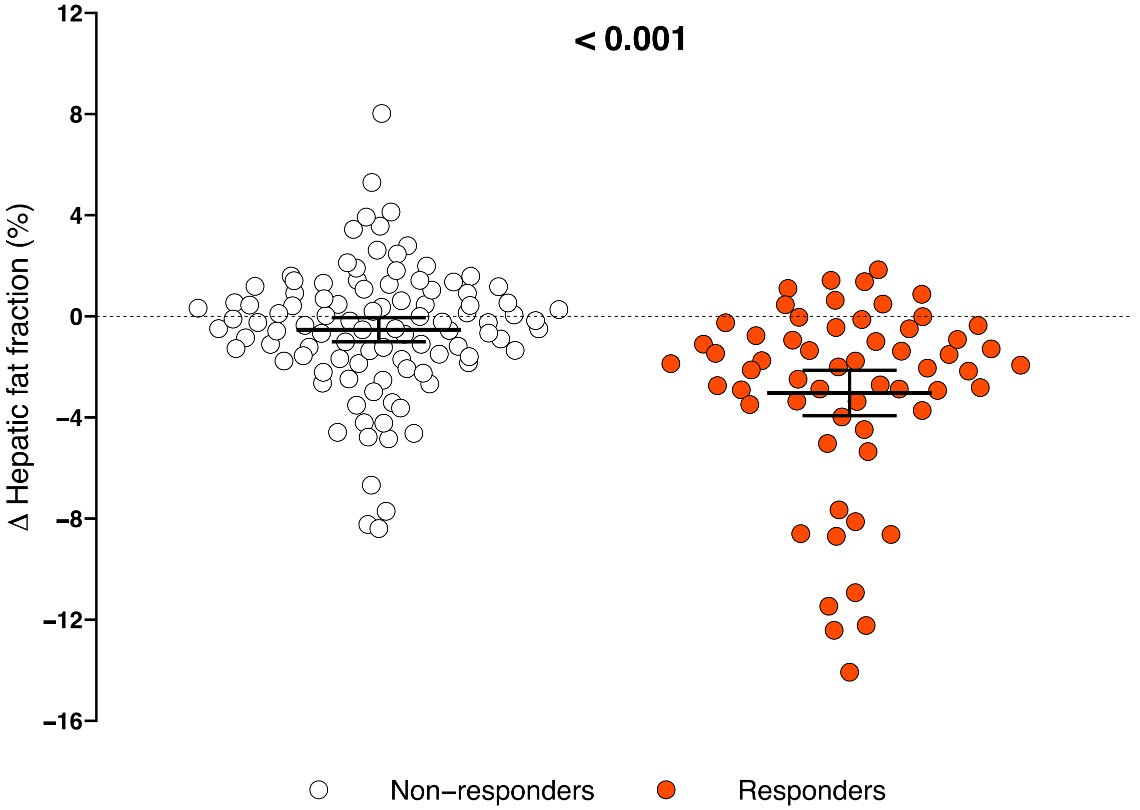

**Figure S8.** Changes in hepatic fat fraction in participants who did not achieve clinically significant body weight loss (<5%, namely non-responders) compared to those who achieved a clinically significant body weight loss (≥5%, namely responders). Data are raw means with 95% confidence interval. Between-group differences (non-responders vs. responders) were evaluated using linear mixed-effects models with repeated measures adjusted for sex. *P* value was corrected using the Benjamini-Hochberg False Discovery Rate (FDR) method to control for multiple comparisons across outcomes. Statistically significant differences are displayed in bold (FDR-adjusted *P* < 0.05). Hepatic fat fraction was assessed by magnetic resonance imaging. Changes were calculated as postintervention minus preintervention values.

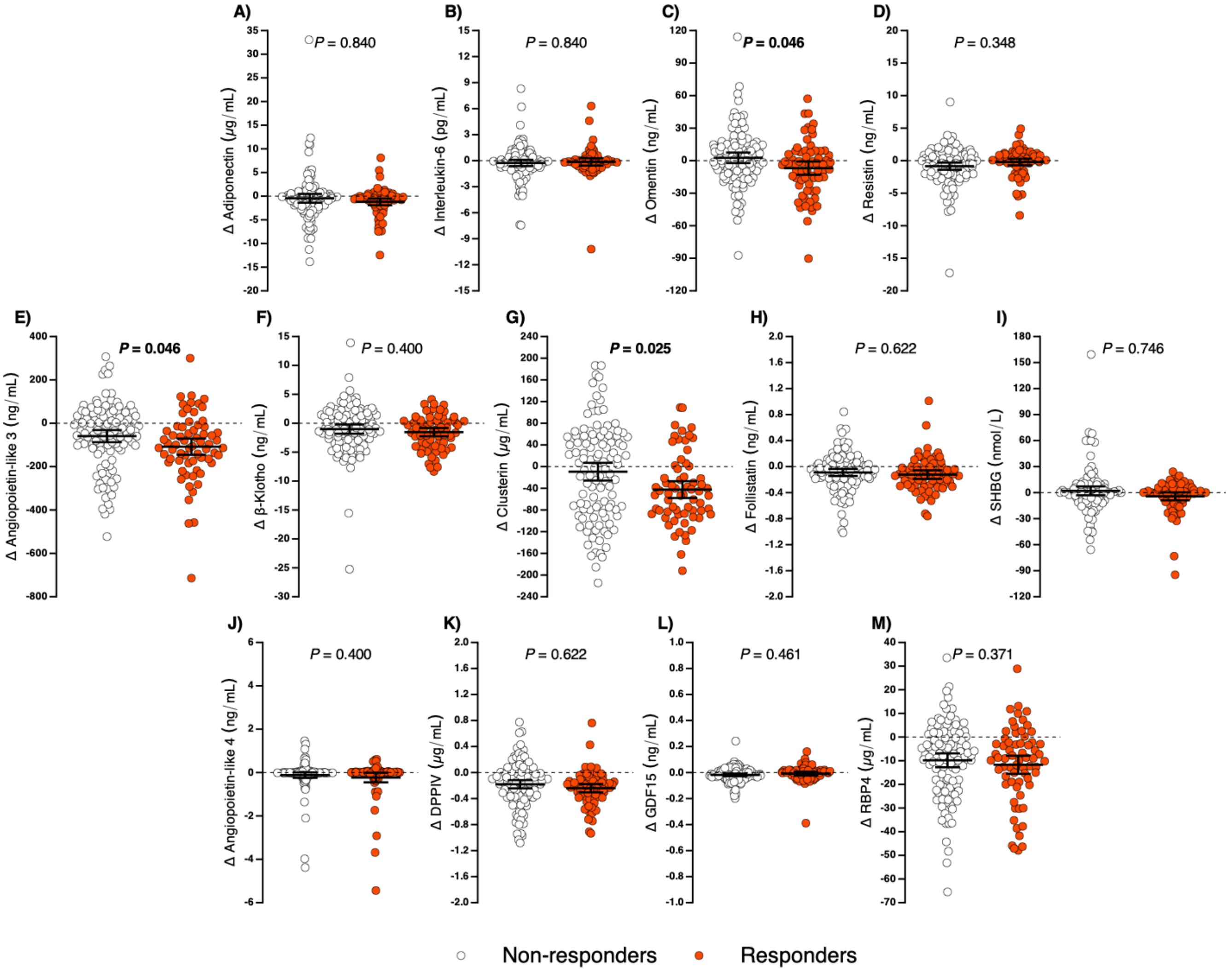

**Figure S9.** Changes in adiponectin (A), interleukin-6 (B), omentin (C), resistin (D), angiopoietin-like 3 (E), β-Klotho (F), clusterin (G), follistatin (H), sex hormone-binding globulin (SHBG; I), angiopoietin-like 4 (J), dipeptidyl peptidase IV (DPPIV; K), growth differentiation factor 15 (GDF15; L), and retinol-binding protein 4 (RBP4; M) in participants who did not achieve clinically significant body weight loss (<5%, namely non-responders) compared to those who achieved a clinically significant body weight loss (≥5%, namely responders). Data are raw means with 95% confidence interval. Between-group differences (non-responders vs. responders) were evaluated using linear mixed-effects models with repeated measures adjusted for sex. *P* values were corrected using the Benjamini-Hochberg False Discovery Rate (FDR) method to control for multiple comparisons across outcomes. Statistically significant differences are displayed in bold (FDR-adjusted $P < 0.05$). Adipokines, hepatokines, and adipo-hepatokines were log10-transformed for statistical analyses; however, results are presented in the original units for improved interpretability. Changes were calculated as postintervention minus preintervention values.

**Table S5.** CONSORT 2025 checklist of information to include when reporting a randomized trial.

| Section/Topic | Item No | CONSORT 2025 checklist item description | Reported on page No |
|---|---|---|---|
| | | **Title and abstract** | |
| Title and structured abstract | 1a | Identification as a randomized trial | 1 |
| | 1b | Structured summary of the trial design, methods, results, and conclusions | 4 |
| Trial registration | 2 | Name of trial registry, identifying number (with URL) and date of registration | 7 |
| Protocol and statistical analysis plan | 3 | Where the trial protocol and statistical analysis plan can be accessed | 7 |
| Data sharing | 4 | Where and how the individual de-identified participant data (including data dictionary), statistical code and any other materials can be accessed | 27 |
| Funding and conflicts of interest | 5a | Sources of funding and other support (eg, supply of drugs), and role of funders in the design, conduct, analysis and reporting of the trial | 27-28 |
| | 5b | Financial and other conflicts of interest of the manuscript authors | 28 |
| Background and rationale | 6 | Scientific background and rationale | 5-6 |
| Objectives | 7 | Specific objectives related to benefits and harms | 6 |
| Patient and public involvement | 8 | Details of patient or public involvement in the design, conduct and reporting of the trial | 7-8 |
| Trial design | 9 | Description of trial design including type of trial (eg, parallel group or crossover), allocation ratio, and framework (for example, superiority, equivalence, non-inferiority or exploratory) | 7-8 |
| Changes to trial protocol | 10 | Important changes to the trial after it commenced including any outcomes or analyses that were not prespecified, with reason | - |
| Trial setting | 11 | Settings (such as community or hospital) and locations (eg, countries or sites) where the trial was conducted | 7-8 |
| Eligibility criteria | 12a | Eligibility criteria for participants | 7 |
| | 12b | If applicable, eligibility criteria for sites and for individuals delivering the interventions (eg, surgeons or physiotherapists) | - |
| Intervention and comparator | 13 | Intervention and comparator with sufficient details to allow replication. If relevant, where additional materials describing the intervention and comparator (eg, intervention manual) can be accessed | 12-13 |
| Outcomes | 14 | Prespecified primary and secondary outcomes, including the specific measurement variable (eg, systolic blood pressure), analysis metric (for example, change from baseline, final value, time to event), method of aggregation (eg, median, proportion), and time point for each outcome | 8-12 |
| Harms | 15 | How harms were defined and assessed (eg, systematically or non-systematically) | - |
| Sample size | 16a | How sample size was determined, including all assumptions supporting the sample size calculation | 13 |
| | 16b | Explanation of any interim analyses and stopping guidelines | - |
| Randomization: | | | |
| Sequence generation | 17a | Who generated the random allocation sequence and the method used | 8 |
| | 17b | Type of randomization and details of any restriction (eg, stratification, blocking and block size) | 8 |
| Allocation concealment mechanism | 18 | Mechanism used to implement the random allocation sequence (eg, central computer/telephone; sequentially numbered, opaque, sealed containers), describing any steps to conceal the sequence until interventions were assigned | 8 |
| Implementation | 19 | Whether the personnel who enrolled and those who assigned participants to the interventions had access to the random allocation sequence | 8 |
| Blinding | 20a | Who was blinded after assignment to interventions (eg, participants, care providers, outcome assessors, data analysts) | 8 |
| | 20b | If blinded, how blinding was achieved and description of the similarity of interventions | 8 |
| Statistical methods | 21a | Statistical methods used to compare groups for primary and secondary outcomes, including harms | 13-15 |
| | 21b | Definition of who is included in each analysis (eg, all randomized participants), and in which group | 16 |
| | 21c | How missing data were handled in the analysis | 13-15 |
| | 21d | Methods for any additional analyses (eg. subgroup and sensitivity analyses), distinguishing prespecified from post hoc | 13-15 |
| Participant flow, including flow diagram | 22a | For each group, the numbers of participants who were randomly assigned, received intended intervention, and were analyzed for the primary outcome | 16 |
| | 22b | For each group, losses and exclusions after randomization, together with reasons | 16 |
| Recruitment | 23a | Dates defining the periods of recruitment and follow-up for outcomes of benefits and harms | 8 |
| | 23b | If relevant, why the trial ended or was stopped | - |
| Intervention and comparator delivery | 24a | Intervention and comparator as they were actually administered (eg, where appropriate, who delivered the intervention/comparator, how participants adhered, whether they were delivered as intended (fidelity)) | 12-13 & 19 |
| | 24b | Concomitant care received during the trial for each group | 12-13 |
| Baseline data | 25 | A table showing baseline demographic and clinical characteristics for each group | 33-34 |
| Numbers analyzed, outcomes and estimation | 26 | For each primary and secondary outcome, by group:<br>• the number of participants included in the analysis<br>• the number of participants with available data at the outcome time point<br>• result for each group, and the estimated effect size and its precision (such as 95% confidence interval)<br>• for binary outcomes, presentation of both absolute and relative effect size | 16-20 & 33-34 & 42-44 |
| Harms | 27 | All harms or unintended events in each group | 16 |
| Ancillary analyses | 28 | Any other analyses performed, including subgroup and sensitivity analyses, distinguishing pre-specified from post hoc | 13-15 & 17-20 |
| Interpretation | 29 | Interpretation consistent with results, balancing benefits and harms, and considering other relevant evidence | 21-26 |
| Limitations | 30 | Trial limitations, addressing sources of potential bias, imprecision, generalisability, and, if relevant, multiplicity of analyses | 24-25 |
